# The genetic drivers of juvenile, young, and early-onset Parkinson’s Disease in India

**DOI:** 10.1101/2023.06.18.23291407

**Authors:** Shan V Andrews, Prashanth L Kukkle, Ramesh Menon, Thenral S Geetha, Vinay Goyal, Rukmini Mridula Kandadai, Hrishikesh Kumar, Rupam Borgohain, Adreesh Mukherjee, Pettarusp M Wadia, Ravi Yadav, Soaham Desai, Niraj Kumar, Deepika Joshi, Sakthivel Murugan, Atanu Biswas, Pramod K Pal, Merina Oliver, Sandhya Nair, Anbu Kayalvizhi, Praveena L Samson, Manjari Deshmukh, Akshi Bassi, Charugulla Sandeep, Nitin Mandloi, Oliver B Davis, Melissa A Roberts, Dara E Leto, Anastasia G Henry, Gilbert Di Paolo, Uday Muthane, Shymal K Das, Andrew S Peterson, Thomas Sandmann, Ravi Gupta, Vedam L Ramprasad, Parkinson Research Alliance of India (PRAI)

**Affiliations:** Denali Therapeutics, South San Francisco, USA; Manipal Hospital, Miller Road, Bangalore, India; Parkinson’s Disease and Movement Disorders Clinic, Bangalore, India; MedGenome Labs Ltd, Bangalore, India; All India Institute of Medical Sciences (AIIMS), New Delhi, India; Medanta Hospital, New Delhi, India; Medanta, The Medicity, Gurgaon, India; Nizams Institute of Medical Sciences (NIMS), Hyderabad, India; Citi Neuro Centre, Hyderabad, India; Institute of Neurosciences Kolkata, Kolkata, India; Bangur Institute of Neurosciences and Institute of Post Graduate Medical Education and Research (IPGME&R), Kolkata, India; Jaslok Hospital and Research Centre, Mumbai, India; National Institute of Mental Health and Neurosciences (NIMHANS), Bangalore, India; Department of Neurology, Shree Krishna Hospital and Pramukhaswami Medical College, Bhaikaka University, Karamsad, Anand, Gujarat, India; All India Institute of Medical Sciences, Rishikesh, India; All India Institute of Medical Sciences, Bibinagar (Hyderabad Metropolitan Region), India; Department of Neurology, Institute of Medical Sciences, Banaras Hindu University, Varanasi; Parkinson and Ageing Research Foundation, Bangalore, India

**Author notes:** These authors contributed equally to this work (Shan V Andrews, Prashanth L Kukkle, Ramesh Menon). Author is deceased (Shymal K Das). **Correspondence to**: Shan V. Andrews, PhD Denali Therapeutics 161 Oyster Point Blvd. South San Francisco, CA 94080. **Relevant conflicts of interest/financial disclosures**: SVA, OBD, MAR, DEL, AGH, GDP, and TS are full-time employees and shareholders of Denali Therapeutics. RM, TSG, SM, MO, SN, AK, PLS, MD, AB, CS, NM, ASP, RG, and VLR are current or former employees of MedGenome Labs. **Funding**: No funding was received towards this work.

## Abstract

**Background:** Recent studies have advanced our understanding of the genetic drivers of Parkinson’s Disease (PD). Rare variants in more than 20 genes are considered causal for PD, and the latest PD GWAS study identified 90 independent risk loci. However, there remains a gap in our understanding of PD genetics outside of the European populations in which the vast majority of these studies were focused.

**Objectives:** To identify genetic risk factors for PD in a South Asian population.

**Methods:** 674 PD subjects predominantly with age of onset ≤ 50 years (encompassing juvenile, young, or early-onset PD) were recruited from 10 specialty movement disorder centers across India over a 2-year period. 1,376 control subjects were selected from the reference population GenomeAsia, Phase 2. We performed various case-only and case-control genetic analyses for PD diagnosis and age of onset.

**Results:** A genome-wide significant signal for PD diagnosis was identified in the *SNCA* region, strongly colocalizing with *SNCA* region signal from European PD GWAS. PD cases with pathogenic mutations in PD genes exhibited, on average, lower PD polygenic risk scores than PD cases lacking any PD gene mutations. Gene burden studies of rare, predicted deleterious variants identified *BSN*, encoding the presynaptic protein Bassoon that has been previously associated with neurodegenerative disease.

**Conclusions:** This study constitutes the largest genetic investigation of PD in a South Asian population to date. Future work should seek to expand sample numbers in this population to enable improved statistical power to detect PD genes in this understudied group.

## Introduction

Parkinson’s disease (PD) is a progressive movement disorder characterized by dopaminergic neuron loss in the substantia nigra. Aging, environmental factors, and genetics contribute to PD risk and progression^1^. To date, common variants in 90 independent risk loci, discovered via genome-wide association studies (GWAS), and rare variants in more than 20 genes have been associated with PD^2,3^. However, most genetic studies of PD have been conducted in individuals of European ancestry^4^. Recent large-scale PD genetics studies in Latinos^5^ and East Asians^6,7^ have both confirmed the cross-ancestry presence of well-established European PD GWAS signals (*i.e.* in the *SNCA* region) and also have provided evidence for the existence of ancestry-specific genetic drivers of PD. However, South Asian populations remain particularly underrepresented in PD genetics studies^4^ and in human genetics studies generally, comprising ∼25% of the world population but only ∼1% of individuals assessed overall in GWAS (as of 2018)^8^. Previous genetics studies of PD in South Asians have been limited by small sample size, a lack of genome-wide interrogation, and/or case-only analyses^9–17^. The success of recent efforts to broaden PD genetic discovery beyond European populations^5–7^, and the high rates of founder effects and consanguinity in South Asians (themselves imparting distinct advantages for variant and gene discovery)^18,19^ render expanded genetics efforts in South Asians a promising opportunity to better understand the genetic basis of PD.

Focusing on individuals with young-(YOPD; typically defined as age of onset, or AoO, ≤ 40) or early-onset (EOPD; 40 < AoO ≤ 50) forms of PD, impart a greater probability of success, because younger age of onset is associated with a greater genetic contribution to disease^3,20^. These results are also relevant for late-onset PD (LOPD; defined herein as AoO > 50) wherever rarer variants associated with earlier-onset or more monogenic PD overlap signals detected via GWAS studies of common variants in LOPD. Several such “pleomorphic” risk loci/genes are known already: *SNCA*, *GBA1*, *LRRK2*, and *VPS13C*^3,21^. Previous genetic studies of early-onset PD have been conducted in a wide variety of ancestral populations, including European^22–26^, East Asian^20,27–35^, and South Asian^12,16,17^ groups. However, most of these early-onset studies were limited by the same factors that have hindered South Asian PD genetics studies generally: small sample sizes, analysis of only a small number of previously known PD genes, and/or lack of control populations to allow for genetic association studies.

In the present study, we address these limitations by conducting the largest genetic study of PD in a South Asian population to date, including genome-wide and case-control discovery approaches. Specifically, we jointly profiled 674 PD subjects predominantly with earlier onset PD from the Parkinson’s Research Alliance of India (PRAI) cohort via genotype array and whole exome sequencing (WES), expanding upon our previous study of 99 subjects from this cohort^17^. Combining these PD data with 1,376 ancestry-matched controls profiled via whole genome sequencing (WGS) and derived from a novel South Asian reference population^18,19^, we performed several studies quantifying variants and genes associated with PD diagnosis and age of onset, utilizing both common and rare variants.

## Methods

674 subjects with PD were recruited through a network of 10 specialty movement disorder centers and/or neurology clinics located across India. PD diagnosis was made via modified UK Brain Bank criteria, and the study was approved by the Ethics Committee of Vikram Hospital (Bengaluru, India) Private Limited; additional details can be found in the previous clinical characterization of this cohort^36^. 1,376 ancestry-matched controls with available WGS data were selected from the GenomeAsia, Phase 2 (GAsPh2) reference cohort^18,19^.

Common variants were profiled with the South Asian Research Genotyping Array for Medicine (SARGAM)^19^, imputed with 6,461 samples from a GAsPh2 reference panel^18,19^, and subjected to stringent quality control (see Supporting Information Methods). Several steps were taken to identify and address genotype measurement error stemming from use of the first iteration of the SARGAM platform (see Supporting Information Methods). A GWAS of PD risk was performed for 4,499,703 common variants (study minor allele frequency or MAF > 5%) by comparing sporadic PD cases (N = 515) to matched controls (N = 1,363), adjusting for sex, age, and the first 10 principal components to account for population stratification. A second association analysis of PD AoO (N = 484 sporadic PD cases) was performed similarly. A significance level of P < 5 × 10^−8^ was used to define genome-wide significance.

WES data was generated for 576 subjects with PD. Variants were called with Sentieon’s GATK best practices germline pipeline and combined with results from WGS data for an additional 92 cases. Using American College of Medical Genetics (ACMG) criteria, we identified pathogenic, likely pathogenic, or variants of uncertain significance (VUS) in known PD genes in these 668 cases, as described previously (see Supporting Information Methods)^17^. For one identified variant in *GBA1*, p.Ser164Arg, we compared levels and activity^37^ of the corresponding enzyme β-Glucocerebrosidase (GCase) in homozygous variant knock-in (KI), *GBA1* knock-out (KO), and wild-type (WT) A549 cells (see Supporting Information Methods).

We established an ancestry-normalized polygenic risk score (PRS) using variant-specific weights from a recent GWAS study of PD in Europeans^2^, and an adjustment procedure to apply this score in a South Asian ancestry sample, as described previously^38^. PRS distributions were compared between cases and controls and within PD cases defined by presence of a PD mutation, with a significance level of P < 0.05.

To perform gene burden studies of rare variants (MAF < 1%) between cases (WES, N = 633) and controls (WGS, N = 1363), we first accounted for case-control platform differences by implementing a previously described calibration procedure informed by synonymous variants (see Supporting Information Methods)^39^. Selected parameters from the calibration procedure were then used in gene burden testing of qualifying variants (QVs) defined based on their predicted functional consequences. Case-control analysis was performed using Fisher’s exact test with multiple testing correction applied across all tests (see Supporting Information Methods).

## Data sharing

Summary statistics for PD diagnosis GWAS and PD age of onset GWAS are available on Zenodo (https://doi.org/10.5281/zenodo.8436983). Analysis code is also available on Zenodo (https://doi.org/10.5281/zenodo.10011733). Summary statistics for the gene burden analyses are available in the supplementary tables. Access to GAsPh2 data can be obtained through https://browser.genomeasia100k.org/#tid=access. Access to European PD GWAS summary statistics including the 23andme discovery dataset can be obtained through https://research.23andme.com/collaborate/. Chromatin interaction data from CNS cell types can be obtained through https://genome.ucsc.edu/s/nottalexi/glassLab_BrainCellTypes_hg19.

## Results

### Data and Analysis Overview

Genetic data was available on a total of 674 PD subjects, encompassing sporadic PD (N = 532; 78.9%) and familial PD (N = 142; 21.1%). Of these 674 subjects, genotype array data was generated on 659 subjects. WGS data was available for a pilot cohort of 92 subjects, as described previously^17^, and we generated WES data on 576 subjects. Control subjects were selected from available WGS from GAsPh2; a total of 1,376 subjects met criteria for ancestry matching (**Fig. 1**, Fig. S1). AoO was available for 651 PD subjects (Table S1), and used to assign them to diagnostic groups, as previously described^36^: 23 juvenile-onset PD (JOPD; AoO ≤ 20 years, 3.5%), 313 YOPD (20 < AoO ≤ 40, 48.1%) and 289 EOPD (40 < AoO ≤ 50, 44.4%) and 26 LOPD (AoO > 50, 4.0%). Case and control genetic data were combined to define genetic drivers of PD, encompassing common variant-(minor allele frequency, MAF > 5%) and/or rare variant (MAF < 1%) - based studies (**Fig. 1**).

**Figure 1.**
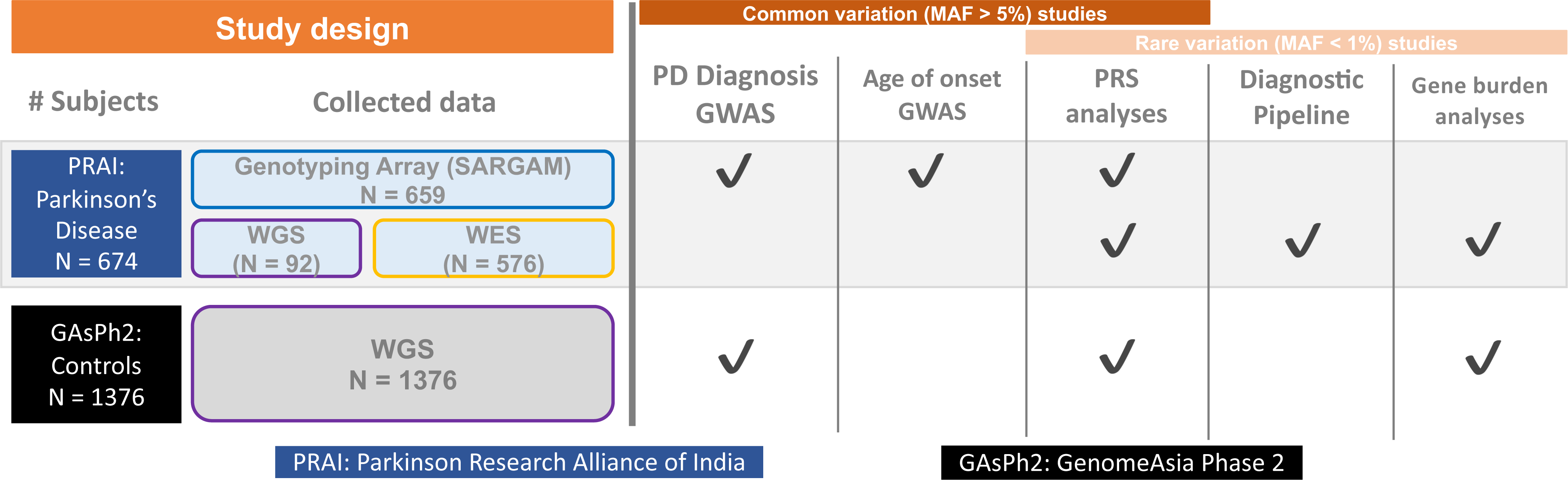
Study overview. A total of 674 PD patients, derived from the PRAI cohort, had some form of genetic data. Of this group, 659 patients had available SARGAM (genotyping array) data. NGS data consisted of 92 subjects with WGS data^17^ and 576 subjects with WES data, which were merged together to effectively create WES data on 668 samples (see Materials and Methods). A total of 1,376 control subjects were used after ancestry-matched to PD cases; these subjects were derived from WGS samples from GAsPh2. 5 major analyses were completed, each of which included common (MAF > 5%) and/or rare variants (MAF < 1%) and different genetic data types from cases and/or controls.

### A genome-wide significant signal for PD diagnosis in the *SNCA* region mirrors signal in Europeans

After implementing a procedure to identify and correct genotyping errors from the novel SARGAM platform (see Supporting Information Methods, Figs. S2-S3), we conducted a GWAS of PD diagnosis using genetic data from sporadic PD samples (N_cases_ = 515) and ancestry matched controls (N_controls_ = 1,363; Fig. S1). Two loci reached genome-wide significance, including one containing the canonical PD gene *SNCA* (**Fig. 2A**). The lead *SNCA* region SNP in both the PRAI cohort and the latest GWAS of PD diagnosis in European samples^2^, rs356182 (p_PRAI_ = 4.11E-11), is harbored in a neuronal enhancer element that physically interacts with the *SNCA* promoter. This enhancer-promoter loop is not observed in the other cell types (microglia and oligodendrocytes) assayed for chromatin interactions in the same dataset^40^ (**Fig. 2B**). The PRAI PD and European PD *SNCA* signals are highly colocalized (coloc^41^ posterior probability = 1; **Fig. 2C**) and show a consistent direction of effect (**Fig. 2D**). The other genome-wide significant locus harbored a lead SNP (rs59330234; p = 1.02E-8) in an intron of *CCDC85A* and has not been previously observed in European PD GWAS. Overall, there was very little concordance genome-wide between PRAI PD GWAS and European PD GWAS signal (excluding the *SNCA* region), even when considering loci reaching a suggestive (p < 1E–5) significance threshold in the PRAI PD GWAS (Fig. S4). In our (case-only) GWAS of AoO in sporadic PD samples (N_cases_ = 484), no genome-wide significant signals were observed (Figs. S2 and S5). Summary statistics for both the PD diagnosis and age of onset GWAS analyses are available (see Data Sharing).

**Figure 2.**
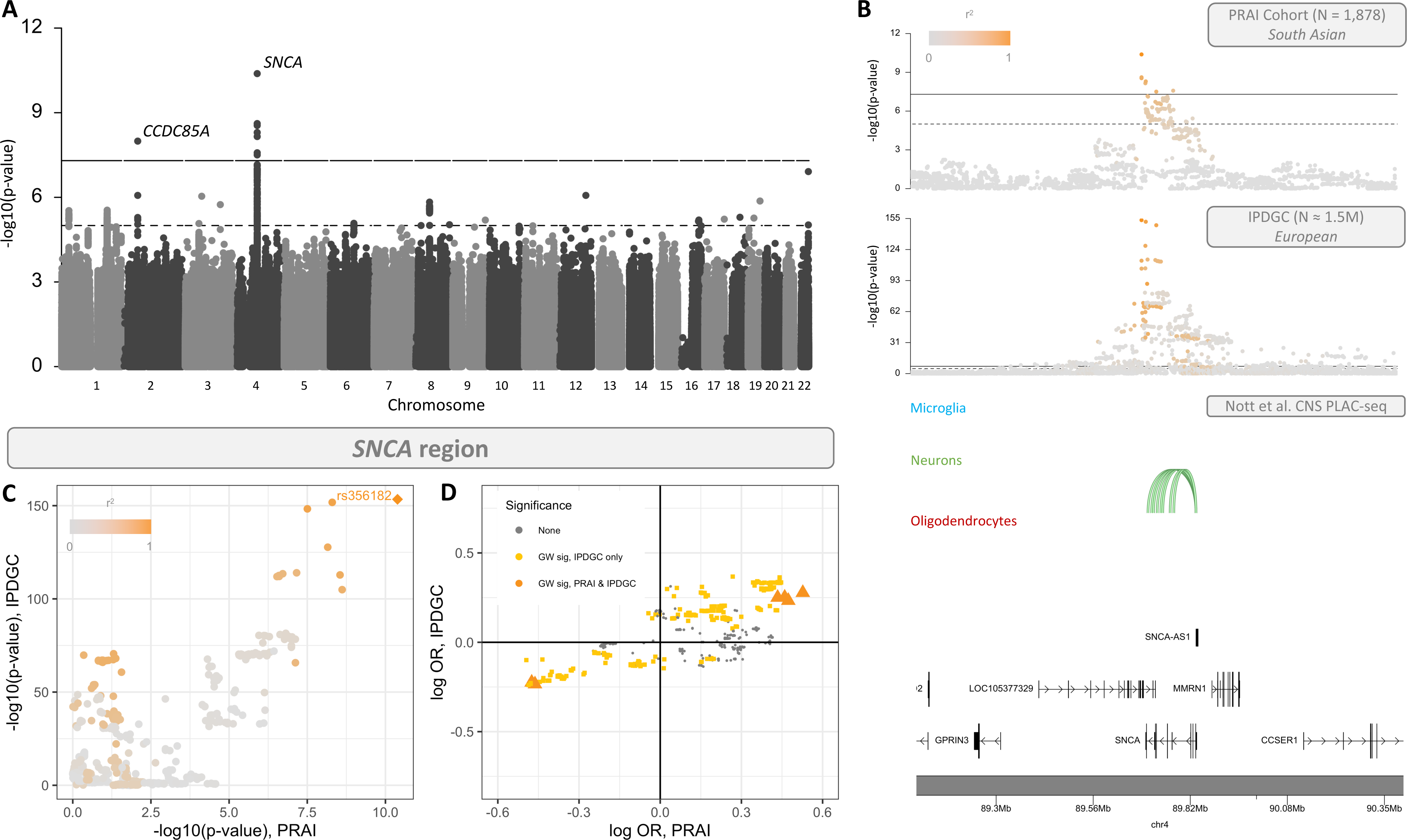
PD Diagnosis GWAS results and concordance of *SNCA* region signals in South Asian and European populations. **(A)** Manhattan plot for PD diagnosis in the joint PRAI (PD cases) - GAsPh2 (controls) cohort for PD diagnosis. Lighter dashed line indicates suggestive significance (p = 1E-5) and darker dashed line indicates genome-wide significance (p = 5E-8). **(B)** Regional association plots in *SNCA* region for South Asian cohort in current study (top panel) and Europeans (International Parkinson’s Disease Genomics Consortium, “IPDGC”, second panel from top) from Nalls et al^2^. Points are colored by r^2^ value in study cohort and 1000 Genomes European samples, respectively. Also plotted are chromatin interaction loops identified in microglia, neurons, and oligodendrocytes^40^ in this region and overlapping a SNP reaching suggestive significance from the current study. **(C) + (D)** Comparison of PRAI-GAsPh2 cohort and SNCA region signals via p-value **(C)** and effect size and direction (via log odds ratio**; D)**.

### *PRKN* variants are a major driver of PD across diagnostic groups

We combined WGS data from a pilot cohort of 92 subjects (previously described^17^) with newly generated WES data for 576 additional individuals (Fig. S6) and assessed them for the presence of pathogenic, likely pathogenic, risk variants, or VUS in a list of 64 previously-associated PD genes by applying ACMG criteria (see Supporting Information Methods and Figs. S7-S8). Among the 649 subjects with available AoO information, pathogenic or likely pathogenic variants (Table S2) were more frequently observed in cases with an earlier age of onset, ranging from 47.6% of JOPD cases (10 of 21 cases), 10.2% of YOPD cases (32 of 313), 3.8% of EOPD cases (11 of 289), and none in LOPD cases (0 of 26; **Fig. 3A**). The total diagnostic yield, defined as the percentage of cases having a pathogenic or likely pathogenic variant in a PD gene, was 8.1%. Of the 33 J/Y/EOPD cases with a pathogenic variant, 22 cases harbored a *PRKN* mutation (**Fig. 3B**). Almost all pathogenic *PRKN* variants were homozygous deletions (20/22 cases) which were concentrated in the 3rd and 4th exons of *PRKN* (Fig. S9). No clinical features displayed strong differences between *PRKN* pathogenic variant carriers and non-carriers (Table S3). Pathogenic mutations were also observed in *PINK1* (N_cases_ = 4), *CHCHD2* and *VPS13C* (both N_cases_ = 2), and *ATP13A2*, *PLA2G6*, and *PRRT2* (all N_cases_ = 1; **Fig. 3B**). Likely pathogenic mutations were observed in *PLA2G6* (N_cases_ = 6), *PRKN* (N_cases_ = 6), *CHCHD2* and *MAPT* (both N_cases_ = 2), and *GCH1*, *PINK1*, *SYNJ1*, and *WDR45* (all N_cases_ = 1; Table S4). Next, we broadened considered variants to include risk variants and VUS, and tabulated occurrences of these variants specifically in a list of 21 genes previously described as causal for PD^3^. *GBA1* mutations were the most frequent in the cohort (**Fig. 3C**), including variants observed across multiple individuals, all in heterozygous form (Table S4), e.g. p.Ser164Arg (N_cases_ = 10), p.Asp448His (N_cases_ = 6), p.Arg170His, p.Ser276Phe, and p.Ala495Pro (all N_cases_ = 3), and p.Arg434Cys (N_cases_ = 2). *GBA1* p.Ser164Arg was observed in the cohort with a high frequency, but had not been functionally characterized before. We investigated the impact of this variant in a human cell culture model (see Supporting Information Methods). Homozygous p.Ser164Arg KI A549 cells exhibited significantly reduced GCase levels (**Fig. 3D**) and activity (**Fig. 3E**) compared to wild-type cells, at magnitudes akin to *GBA1* KO cells. A list of all identified variants in J/Y/EOPD cases, across all 64 PD-related genes, including the counts of heterozygous and homozygous carriers in cases and controls (when available) is available as Table S4.

**Figure 3.**
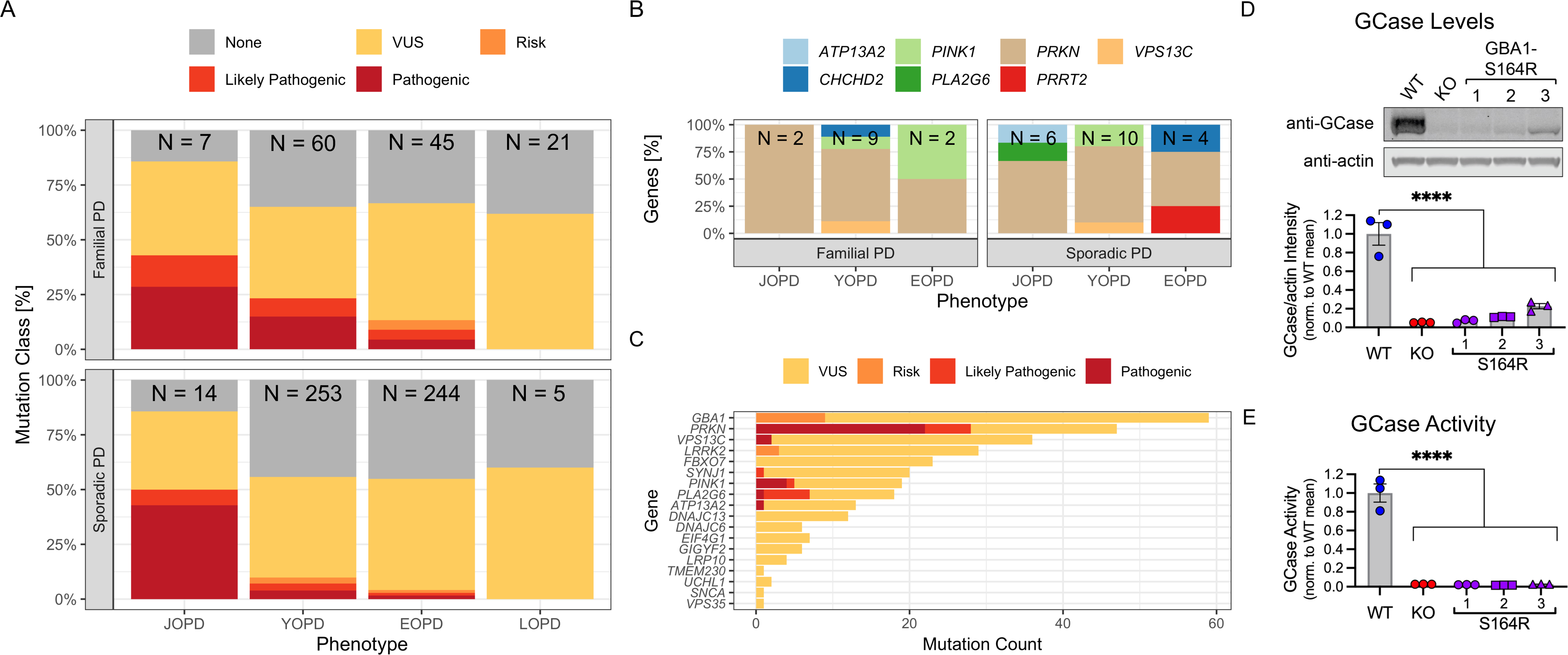
Diagnostic pipeline results. **(A)** Distribution of mutation type in known PD–related genes, by diagnostic group and familial/sporadic PD status. **(B)** Distribution of genes affected by pathogenic variants, by diagnostic group and familial/sporadic PD status. **(C)** Counts of all pathogenic, likely pathogenic, risk, and VUS variants identified in 21 genes previously described as monogenic for PD**. (D)** Representative Western blot and quantification of total cellular GCase levels from A549 *GBA1*-wild type, *GBA1*-KO, and three clonal *GBA1*-S164R variant cell lines. Bars represent mean GCase levels (normalized to beta-actin loading control within each sample) normalized to the mean WT value across all replicates, and points represent values from individual replicates. (Error bars: SEM [*n* = 3]; **** = p<0.0001, ANOVA with Dunnett’s multiple comparisons test). **(E)** Flow cytometry measurement of GCase activity in A549 *GBA1*-wild type, *GBA1*-KO, and three clonal *GBA1*-S164R variant cell lines. Bars represent mean GCase activity normalized to the mean WT value across all replicates, and points represent values from individual replicates. (Error bars: SEM [*n* = 3]; **** = p<0.0001, ANOVA with Dunnett’s multiple comparisons test).

### PD gene pathogenic variant carriers have significantly reduced polygenic burden relative to non-carriers

We next leveraged our imputed SARGAM microarray data to construct a PRS for PD, using weights from the most recent PD GWAS study in European samples^2^, and a previously described method to project a PRS informed by weights of one ancestral population to another^38^. PD cases had a significantly higher PRS score than controls (Wilcoxon test, p-value = 1.94E-11; **Fig. 4A**). To further quantify this difference, we binned samples into deciles (using breakpoints defined in the controls, as previously described^38^) and calculated decile-specific odds ratios for disease, adjusted for age and sex. Membership in the tenth decile was significantly associated with increased odds of disease relative to a joint 5th and 6th decile comparison group^38^ (Odds Ratio = 1.61, p = 0.006; **Fig. 4B**).

**Figure 4.**
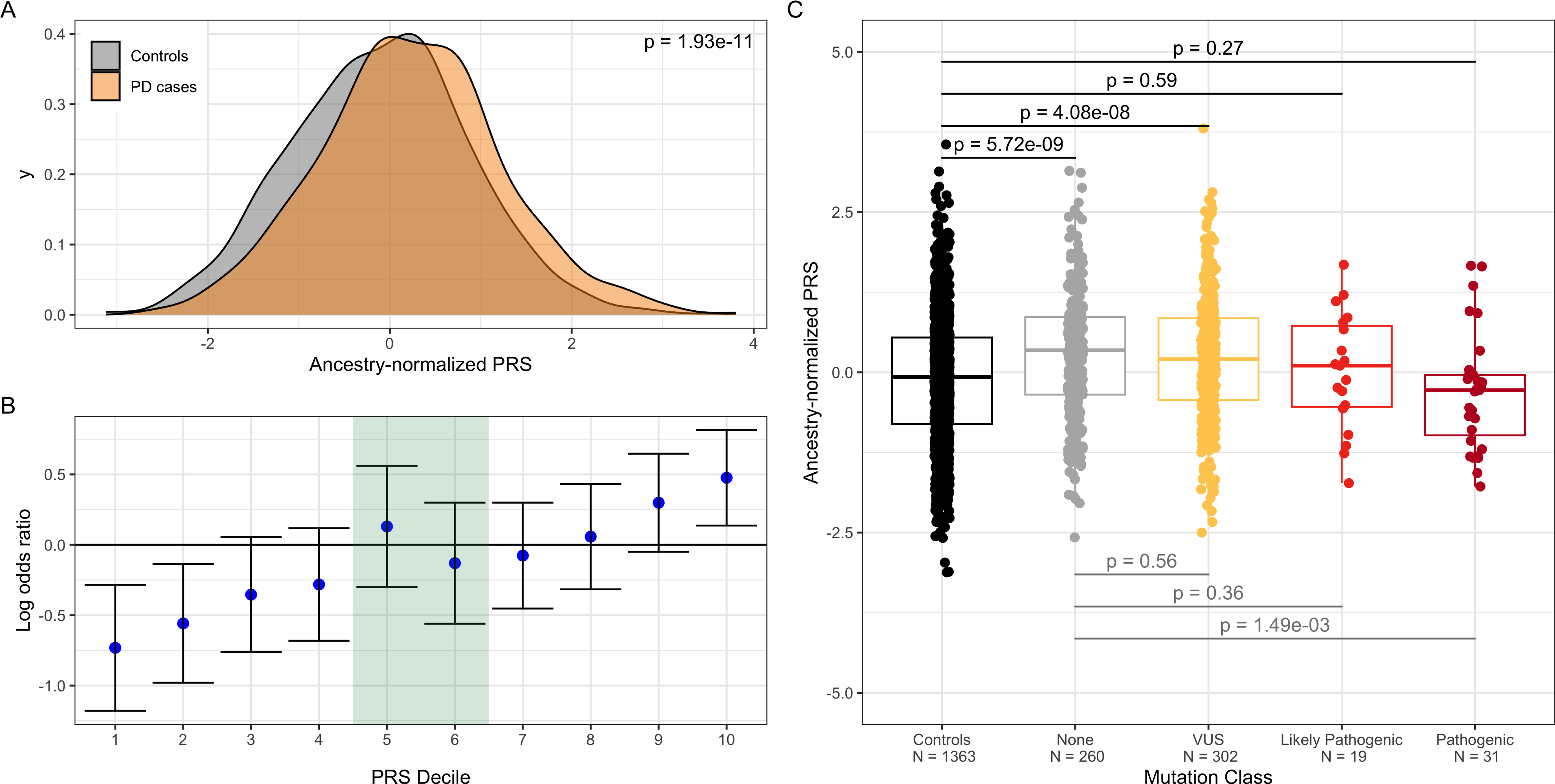
Polygenic risk scores and their integration with diagnostic categories. **(A)** Distributions of ancestry-normalized PRSs in PD cases and controls. **(B)** Odds ratios for disease in PRS-defined deciles in the control group, relative to deciles 5 and 6. **(C)** Distribution of PRSs in controls and diagnostic pipeline-defined categories for PD cases.

Next, we compared the PRS between controls and several groups of PD cases defined by presence of a PD-related mutation (“VUS”, “Likely Pathogenic”, or “Pathogenic”) or lack thereof (“None”), and within those same PD case groups. The “None” (p = 5.72E-9) and “VUS” (p = 4.08E-8) groups exhibited significantly higher PRS than the control group, adjusted for age and sex. Interestingly, the “Pathogenic” group exhibited significantly lower PRS compared to the “None” group (p = 1.49E-3; **Fig. 4C**), adjusted for age and sex. No other comparisons achieved nominal (p < 0.05) statistical significance.

### Gene burden analyses of rare, predicted deleterious variants demonstrate suggestive evidence linking *BSN* to PD

We next performed gene burden analysis with WES data from PD cases (N = 633) and WGS data from controls (N = 1363; Fig. S7). Using a calibration procedure informed by synonymous variants^39^, we derived coverage and variant quality filters to adjust for platform differences between cases and controls (see Supporting Information Methods and Fig. S10). After imposing these filters, and further restricting to variants with MAF < 1% across all ancestral populations, a total of 134,871 variants from the PD case WES data and 195,903 variants from the control WGS data could be evaluated. We selected QVs according to several criteria (see Supporting Information Methods)^41^. For example, the most stringent analysis considered 91 genes with predicted loss-of-function (LoF) variants. Additional, more permissive definitions of QVs, infomed by the *in silico* prediction tools Combined Annotation Depletion-Dependent (CADD)^43^ and Polymorphism Phenotyping v2 (PolyPhen2)^43^ were implemented, along with a minimum carrier count across cases and controls of 5 samples, resulting in different gene counts corresponding to stringency: 1) 2,854 genes carrying LoF + PolyPhen2 “probably damaging” variants, 2) 4,572 genes with LoF + PolyPhen2 “probably damaging” or “possibly damaging” variants, 3) 7,627 genes with variants with CADD score > 20, and 4) 9,209 genes with broadly “Functional Rare” variants (encompassing missense, inframe, frameshift, and nonsense variants) for analysis (**Fig. 5A**). After removing the LoF only tests (low gene count) and gene-QV criterion combinations potentially impacted by confounding (see Supporting Information Methods), a total of 21,412 burden tests were performed, with many genes tested multiple times under different QV criteria (Table S5). No tests surpassed a Bonferroni correction threshold (p = 2.33E-6), likely due to the limited statistical power afforded by the cohort (Fig. S11). However, several genes achieved marginal significance (p < 5E-4) across multiple QV criteria, demonstrating association with PD not entirely restricted to a single set of variants (**Fig. 5B**). For example, we observed suggestive associations for *BSN* under the “Functional Rare” QV criterion (p = 2.57E-5, the most significant result observed overall), but also the LoF + PolyPhen2 “probably damaging” or “possibly damaging” (p = 3.56E-4) and the CADD score > 20 (p = 3.88E-4) criteria. *BSN* encodes the Bassoon protein and is highly expressed in the brain (Gene-Tissue Expression (GTEx) Project v8^45^, Fig. S12A). Interestingly, the variants driving the differential gene burden signal were localized towards the C-terminus (a similar region of the protein implicated in a recent genetics study of progressive supranuclear palsy-like syndrome, or PSP^46^) and were never observed in controls (Fig. S12B).

**Figure 5.**
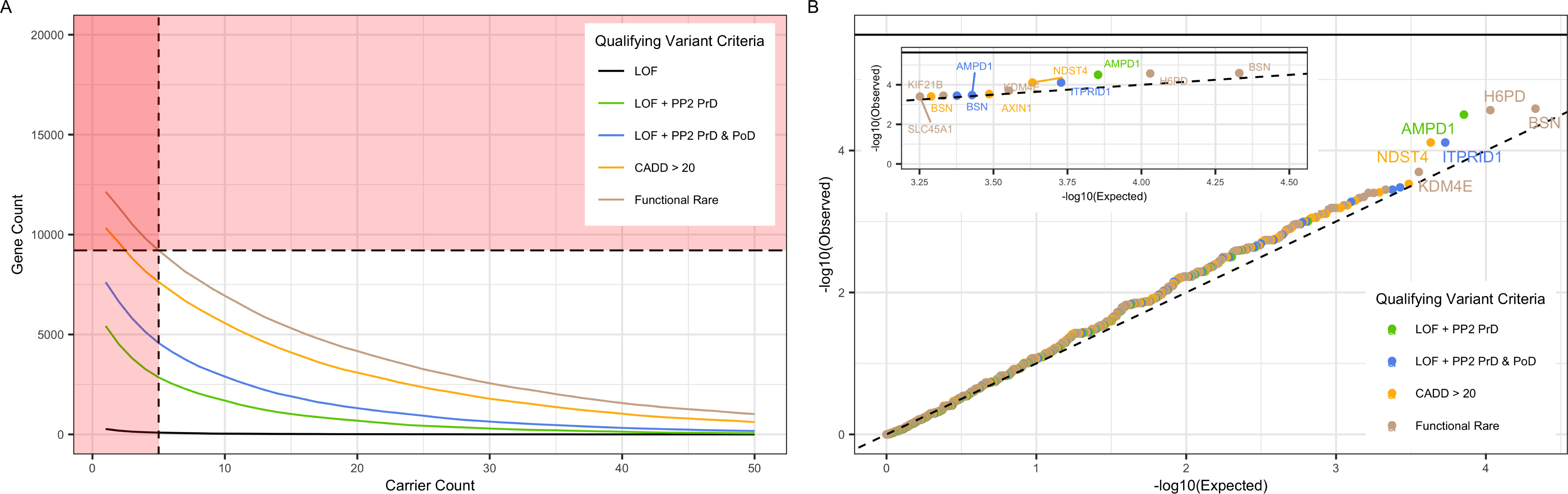
Gene burden analyses of rare variants. **(A)** Number of genes available for analysis as a function of the number of carriers (across cases and controls), for several different definitions of a qualifying variant (QV). Genes with carrier count ≥ 5 were included in gene burden testing. LoF-only analysis not performed due to low number of genes available that met the minimum carrier count threshold. **(B)** QQ-plot displaying results of gene burden studies across QV criteria. Methods and plots adapted from previous gene burden analysis of Alzheimer’s Disease^42^.

## Discussion

We have conducted the largest genetics study of PD in a South Asian population to date and to our knowledge, a study focused on PD patients with an earlier-onset phenotype (AoO ≤ 50 years) and utilizing a diverse set of genetic data types (genotype array and WES in cases, WGS in controls) and analysis approaches (including common and rare variation, variant and gene-level inferences, and PRS). By comparing these PD patients with ancestry-matched controls we identified a genome-wide significant signal in the region of *SNCA*, a canonical PD gene, that recapitulates the European PD GWAS signal from the same region. Approximately 8% of PD patients harbor a pathogenic or likely pathogenic mutation in a previously known PD gene (predominantly, *PRKN*), and carriers of such pathogenic variants have a significantly reduced polygenic burden relative to PD cases without any PD gene mutations. Finally, through case-control comparison of rare variants at the gene level, we demonstrate suggestive evidence for an association of *BSN* and increased PD risk.

The PD diagnosis GWAS yielded two genome-wide significant signals. The first was an intronic SNP in *CCDC85A*, implicating this gene, which is highly expressed in the brain^45^, for the first time in PD. The second signal, in the *SNCA* region, was driven by the lead SNP rs356182, which matches that of one of the two independent *SNCA* region signals reported in a recent European PD GWAS^2^, and has been functionally characterized extensively^47,48^. Neuronal chromatin interaction data^40^ indicates that rs356182 variants impact *SNCA* gene expression in this cell type as shown previously^47,49^, and consistent with the proposed impact of this SNP on upregulating a *SNCA* 5’ untranslated region transcript isoform in frontal cortex^47^. This SNP has also been reported as the lead SNP in *SNCA* region signals in recent PD studies in Latino^5^ and Chinese^7^ populations, and in a multi-ancestry meta-analysis including individuals of European, East Asian, Latin American, and African ancestry^50^. Our results extend the relevance of rs356182 to an additional ancestral population, South Asians, and to earlier onset forms of PD. Beyond this *SNCA* region signal, no regions demonstrating suggestive signal for PD diagnosis (p < 1E-5) in our South Asian cohort showed evidence of association in European samples. Many of these suggestive signals may turn out to be false positives, but some might also correspond to common variation associated with PD risk that is unique to South Asians populations. Future, larger studies of PD in South Asians will be better powered to distinguish these possibilities.

The examination of known PD genes revealed strong rare variant drivers, and the diagnostic yield (∼8%) in our patient population matches that of similar studies in East Asian populations^20,33,34^ (7.9% to 11.6%). Homozygous deletions in *PRKN* comprised the majority of pathogenic variants identified in this cohort, consistent with previous studies demonstrating that homozygous or compound heterozygous *PRKN* variants are the major driver of young- or early-onset PD^51–53^. Furthermore, the identified *PRKN* deletions were concentrated in exons 3 and 4 of *PRKN*, consistent with a recent comprehensive assessment of *PRKN* mutation frequency^54^. We also detected a number of *GBA1* variants in PD cases. However, all were classified via ACMG criteria as being “Risk” or “VUS” variants, and therefore their roles as causal genetic drivers remains to be elucidated. One such *GBA1* variant, p.Ser164Arg, was observed in 10 (unique) PD cases out of 668 (1.5%). A549 cells carrying a homozygous p.Ser164Arg KI displayed a marked decrease in both GCase levels and activity, effectively phenocopying gene KO cells. This result, the first functional evidence for p.Ser164Arg presented to date, is in line with the homozygous occurrence of this variant in Gaucher disease (GD) patients^55–57^, whose GCase levels and/or activity are severely reduced. Data from gnomAD^58^ and a recent comprehensive investigation of *GBA1* variant frequencies^59^ only report occurrence of p.Ser164Arg in South Asians. Along these lines, all previous reports of p.Ser164Arg in GD patients and in PD patients^16^ have come from Indian populations. In the latter instance, p.Ser164Arg was observed in 3 of 250 (1.2%) EOPD patients, a frequency similar to that observed in this cohort. Two additional *GBA1* variants, p.Ser276Phe and p.Arg434Cys, observed in 3 and 2 PD cases respectively, also appear to be only present in South Asian individuals^58,59^. Given this evidence, future functional studies should ascertain if these variants similarly reduce GCase levels and activity as demonstrated here for p.Ser164Arg, and whether they pinpoint novel aspects of *GBA1* biology.

We observed a modestly increased polygenic burden in PD cases relative to controls. When considering rare and common variants jointly, we observed this difference was driven by PD cases lacking a PD gene mutation (“None” group) and those carrying only a VUS. Recent functional studies of induced pluripotent stem cell (iPSC) derived dopaminergic neurons from EOPD patient samples without known PD gene mutations displayed increased α-synuclein accumulation and reduced lysosomal membrane protein abundance compared to control iPSC lines^60^. Our findings are consistent with these results in arguing for the existence of genetic contributions to EOPD beyond rare mutations in known PD risk genes. It was also observed in our study that PD cases with a pathogenic PD gene mutation had significantly reduced polygenic burden relative to PD cases lacking a PD gene mutation. Similar findings of reduced polygenic burden in mutation carrier cases relative to non-carrier cases have also been observed for phenotypes such as autism spectrum disorder^61^, breast and colorectal cancer, diabetes, osteoporosis, and short stature^62^. These observations are consistent with a liability threshold model of disease by which rare, pathogenic variant carriers require a smaller contribution from common variants to reach a disease diagnosis, compared to non-carriers^63^. However, because only a limited number of studies have examined the interplay between rare and common variant genetic drivers in PD specifically^64,65^, this is an important area of future work.

Gene-based burden analyses revealed a suggestive association for *BSN*, which has had limited connection to PD previously. As mentioned above, *BSN* encodes Bassoon, a large synaptic scaffolding protein that plays a critical role in assembling the presynaptic active zone, where synaptic vesicle exocytosis primarily occurs^66^. Bassoon is also a key regulator of presynaptic proteostasis, controlling degradative pathways that have been extensively implicated in PD, such as autophagy, notably in concert with the *bona fide* PD-linked protein Parkin^67^. Recent human genetic and preclinical studies have also directly implicated Bassoon in neurodegenerative diseases, by identifying mutations in individuals with PSP^46^, a primary tauopathy, and showing a role of Bassoon in the seeding and toxicity of tau pathology in mice^68^. Our findings suggest a broader involvement of Bassoon in neurodegenerative disease, and future work should look to evaluate the functional impact of the individual *BSN* variants driving the gene-level association we observed.

Our study has limitations. The total sample size for our study population consisted of roughly 2,000 cases and controls, limiting the statistical power for all variant- and gene-based discovery analyses. Moreover, given this sample size, and the rarity of early-onset PD cases generally, we were not able to derive a suitable validation cohort for our findings. In some instances, our findings are validated by previous studies in other cohorts or populations, such as the *SNCA* region signal for PD diagnosis or high frequency of *PRKN* deletions in our cohort. Other novel findings, such as the reduced polygenic burden observed in PD cases carrying pathogenic variants, or the suggestive association of *BSN*, require replication in additional South Asian PD cohorts and in PD cohorts more generally.

The Global Parkinson’s Genetics Program (GP2) was recently initiated to directly address the limited insights into PD disease etiology and limited potential for equitable personalized medicine due to the strong bias of European ancestry samples in previous PD genetics studies^4,69^. This work is an initial step in addressing this knowledge gap in a South Asian population. With this effort and parallel GP2-fueled PD genetics studies in India^70^, the knowledge of PD genetic risk factors in South Asians, and therefore understanding of PD genetics as whole, should increase substantially in the coming years.

## Acknowledgements

We thank the study participants for their time and contribution of genetic and clinical information to perform this study. We thank J. Fah Sathirapongsasuti for his contribution to the data analysis and study design.

## Authors’ Roles

Design: SVA, PLK, RM, OBD, MAR, DEL, AGH, AP, TS, RG, VLR

Execution: All authors

Analysis: SVA, PLK, RM, TSG, MO, SN, AK, PLS, MD, AB, CS, NM, OBD, MAR, TS, RG

Writing: SVA, PLK, RM, OBD, MAR, GDP, TS, RG

Editing of final version of the manuscript: SVA, PLK, RM, OBD, MAR, DEL, AGH, GDP, TS, RG, VLR

