## Supplementary Figures for "The genetic drivers of juvenile, young, and early-onset Parkinson’s Disease in India"

**Figure S1.** UMAP representation of GAsPh2 reference population along with PD cases and ancestry-matched controls selected from GAsPh2

**Figure S2.** Analysis flowchart for studies of common variants (MAF > 5%)

**Figure S3.** Corrective procedures for SARGAM data and PD diagnosis GWAS analysis.

**Figure S4.** Genome-wide comparison of PRAI (South Asian) cohort PD GWAS signal vs International Parkinson’s Disease Genomics Consortium (IPDGC; European) cohort PD GWAS signal

**Figure S5.** Age of onset GWAS cohort description and Manhattan plot

**Figure S6.** Boxplots for WES data QC metrics

**Figure S7.** Analysis flowchart for studies of rare variants (MAF < 1%)

**Figure S8.** Diagnostic pipeline workflow

**Figure S9.** Number of pathogenic/likely pathogenic deletions affecting different *PRKN* exons

**Figure S10.** A synonymous variant informed calibration procedure corrects platform differences between PD cases (WES data) and GAsPh2 controls (WGS data) for gene burden studies.

**Figure S11.** Power curves for gene burden studies

**Figure S12.** *BSN* gene expression and variants driving differential gene burden result.

**Figure S1. UMAP representation of GAsPh2 reference population along with PD cases and ancestry-matched controls selected from GAsPh2**

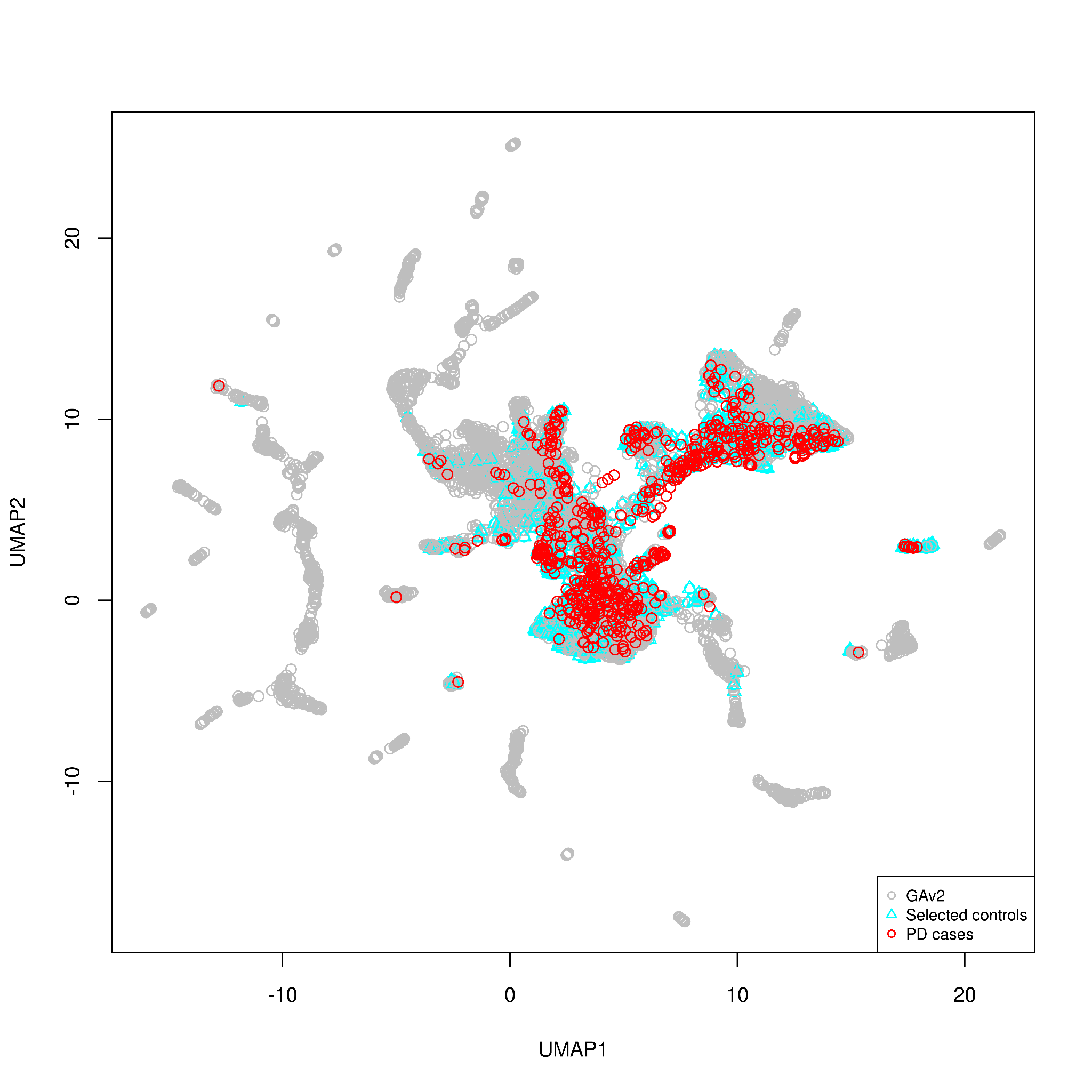

**Figure S2. Analysis flowchart for studies of common variants (MAF > 5%)**

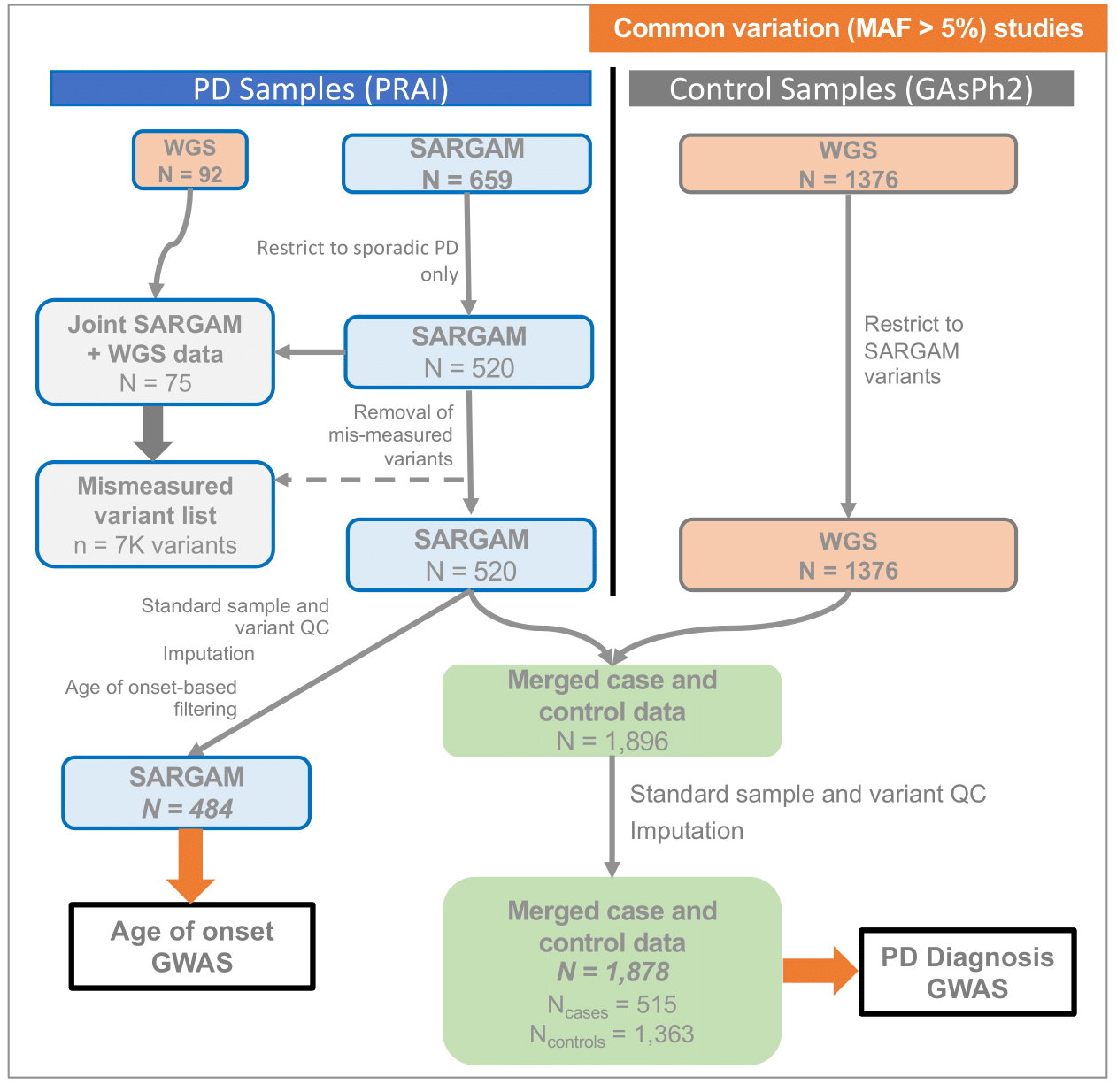

**Figure S3. Corrective procedures for SARGAM data and PD diagnosis GWAS analysis.** A) Relationship of measurement error (defined as absolute value of allele frequency measured via SARGAM platform vs that measured via WGS on the same samples, for N = 75 samples with data on both platforms) to WGS-measured frequency B) Relationship of PD diagnosis GWAS p-value to measurement error before measurement error correction C) Relationship of PD diagnosis GWAS p-value to measurement error after removal of mismeasured variants and reperforming imputation. Measurement error here reflects that from pre-corrected data; presence of GWAS p-values for mismeasured variants indicates they were recovered via (re)imputation process. D) Relationship of GWAS p-value (from post measurement error-corrected analysis) for genome-wide significant SNPs to Beagle imputation quality (DR2) metric. E) Quantile-quantile plot for PD diagnosis GWAS analysis, which was adjusted for age, sex, and population stratification.

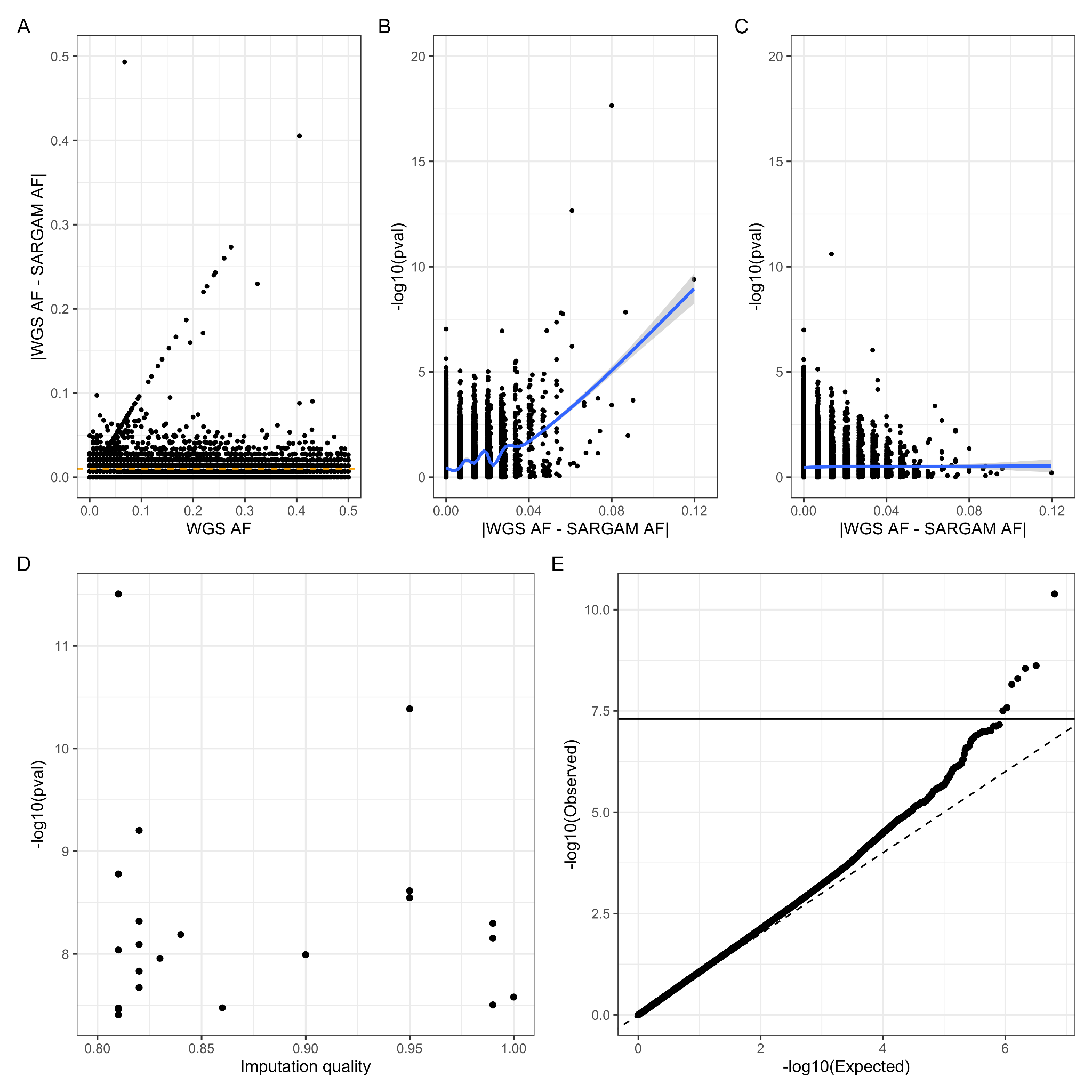

**Figure S4. Genome-wide comparison of PRAI (South Asian) cohort PD GWAS signal vs International Parkinson’s Disease Genomics Consortium (IPDGC; European) cohort PD GWAS signal.** A) PRAI vs IPDGC effect sizes (log odds ratio) at all variants shared between studies (n = 4,114,155); variants in the *SNCA* region are indicated via point shape. B) The same PRAI vs IPDGC comparison, but via -log10(p-value) C) For the lead SNPs in all loci reaching suggestive significance levels (p < 1E-5), the MAF measured in 1000 Genomes SAS samples or EUR samples vs the MAF in the PRAI study cohort. SAS and PRAI MAFs are consistent, as expected, but EUR MAFs are largely smaller than PRAI MAFs. D) For the same suggestive regions in PRAI (excluding the *SNCA* region), the -log10(p-value) vs MAF as seen at the SNP with most significant p-value from the IPDGC European PD GWAS in that region. No strong signal is observed at these SNPs, likely driven by low MAF in Europeans.

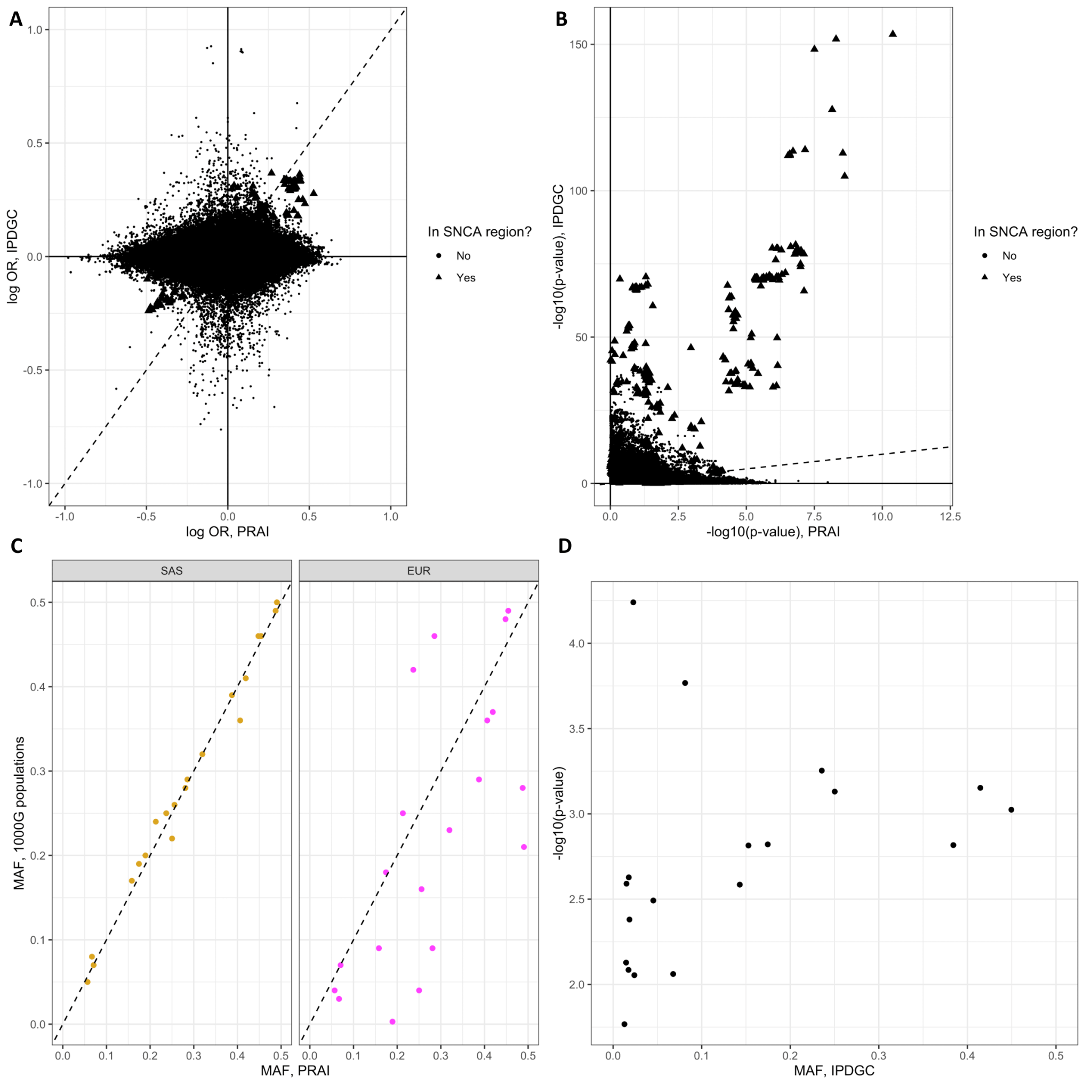

**Figure S5. Age of onset GWAS cohort description and Manhattan plot** A) Histogram of age of onset for full PD case cohort in which variable awas available (N = 651). AoO GWAS was restricted to sporadic PD samples only, those samples with AoO < 50, and removed an outlying sample with AoO = 3. B) Manhattan plot for AoO GWAS, from a model adjusted for sex and population stratification.

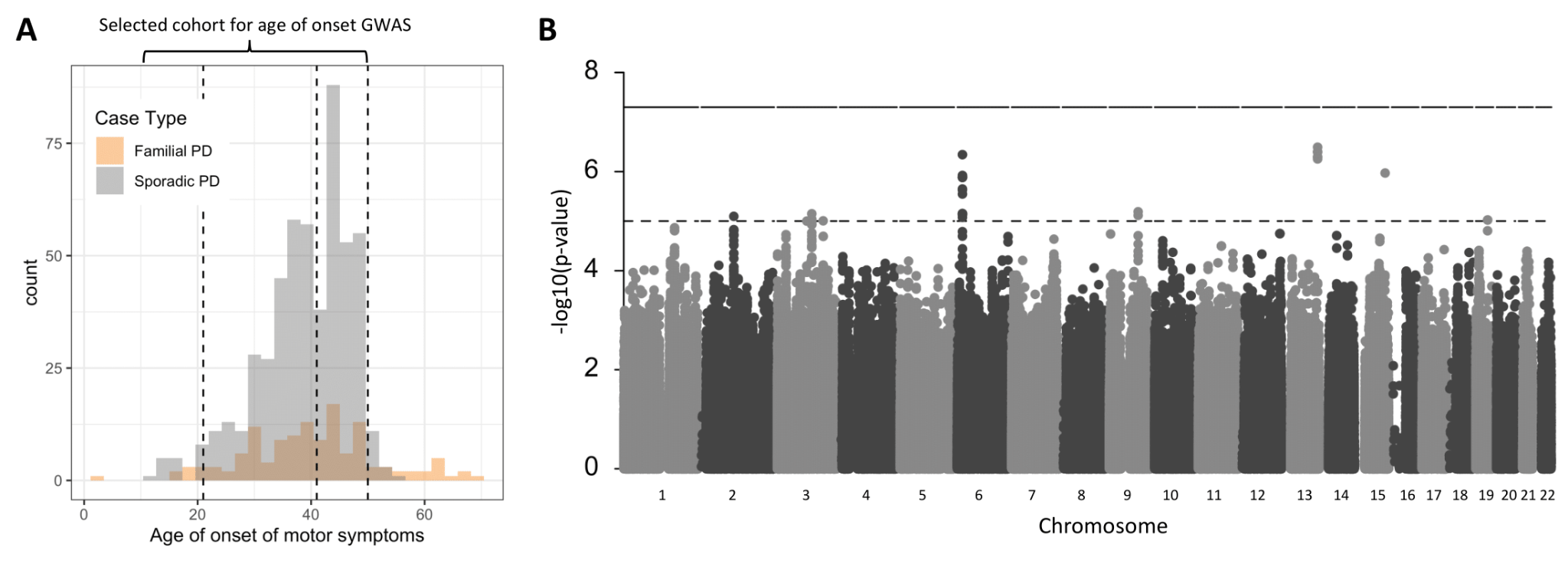

**Figure S6. Boxplots for WES data QC metrics**

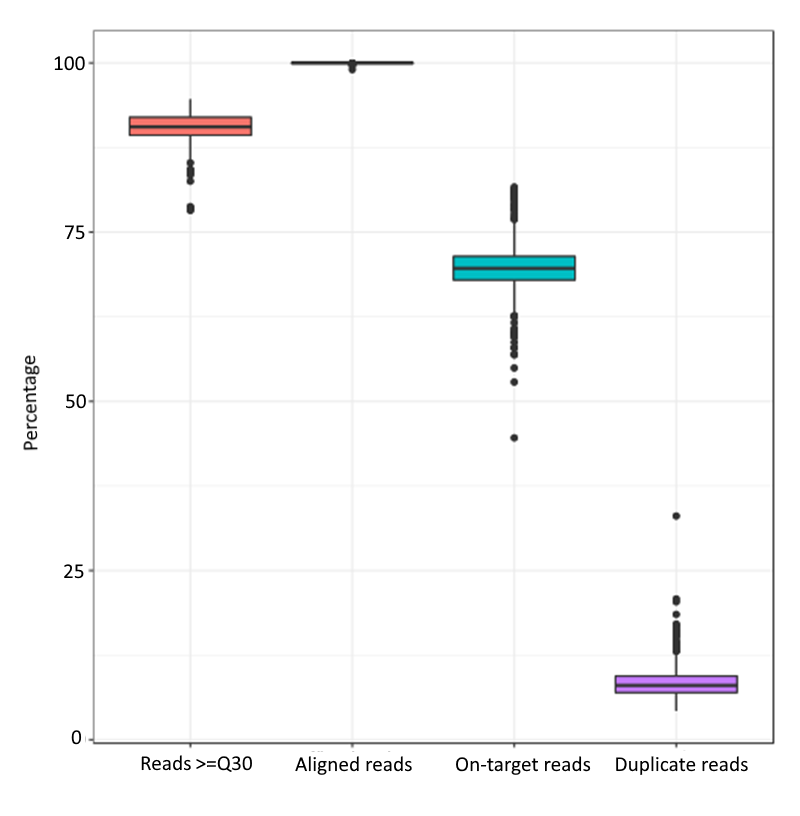

**Figure S7. Analysis flowchart for studies of rare variants (MAF < 1%)**

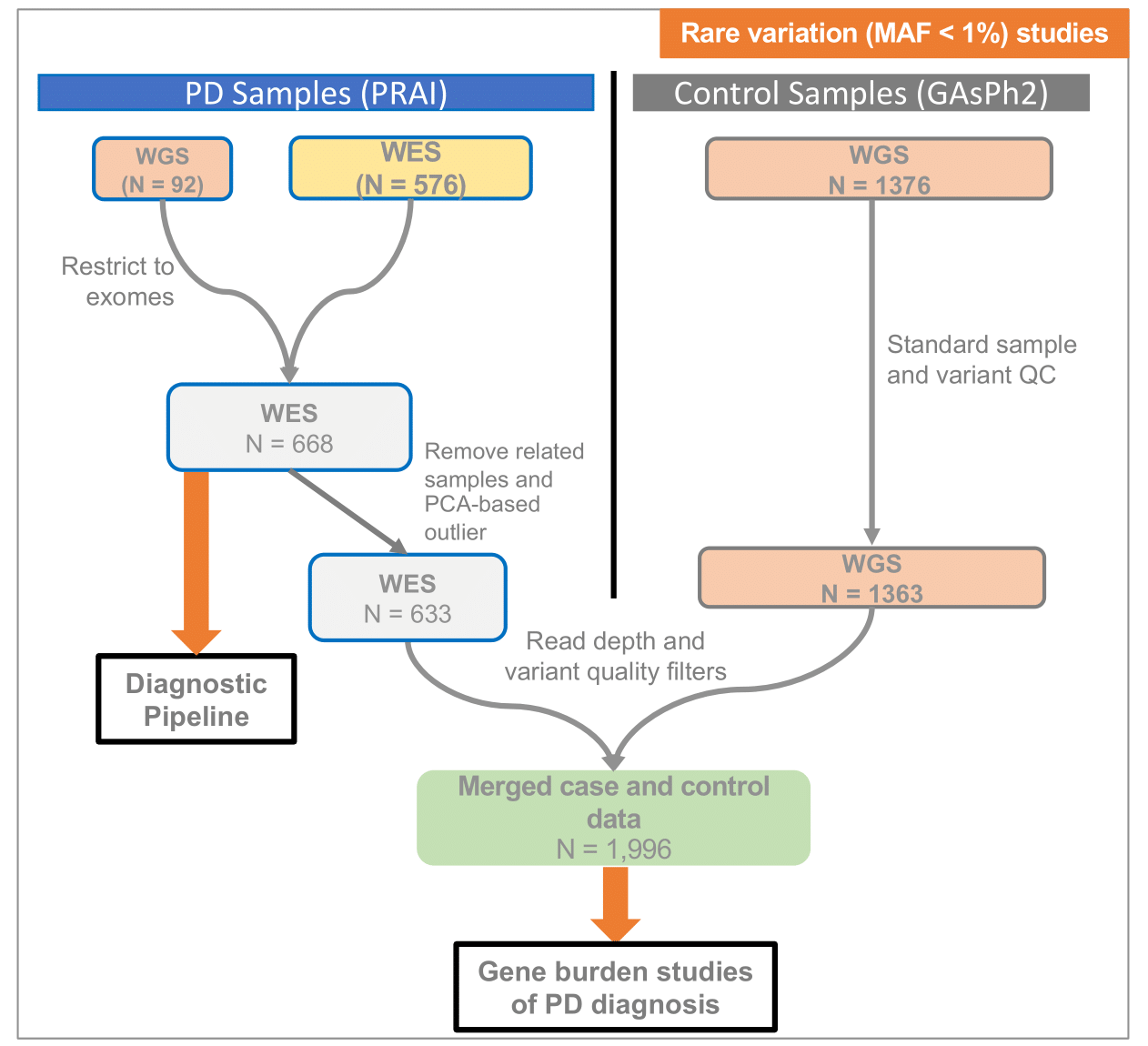

**Figure S8. Diagnostic pipeline workflow**

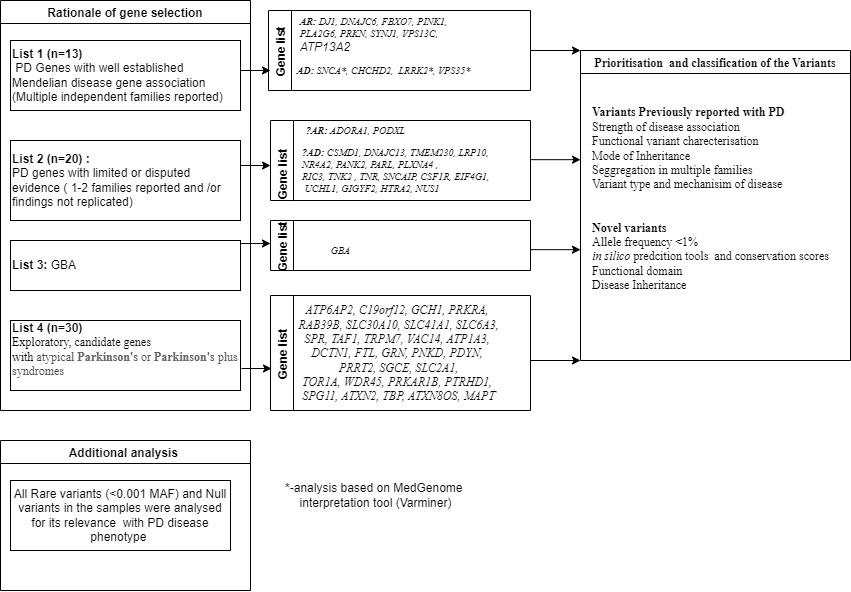

**Figure S9. Number of pathogenic/likely pathogenic deletions affecting different *PRKN* exons.** See accompanying Table S2 for a full list of deletions.

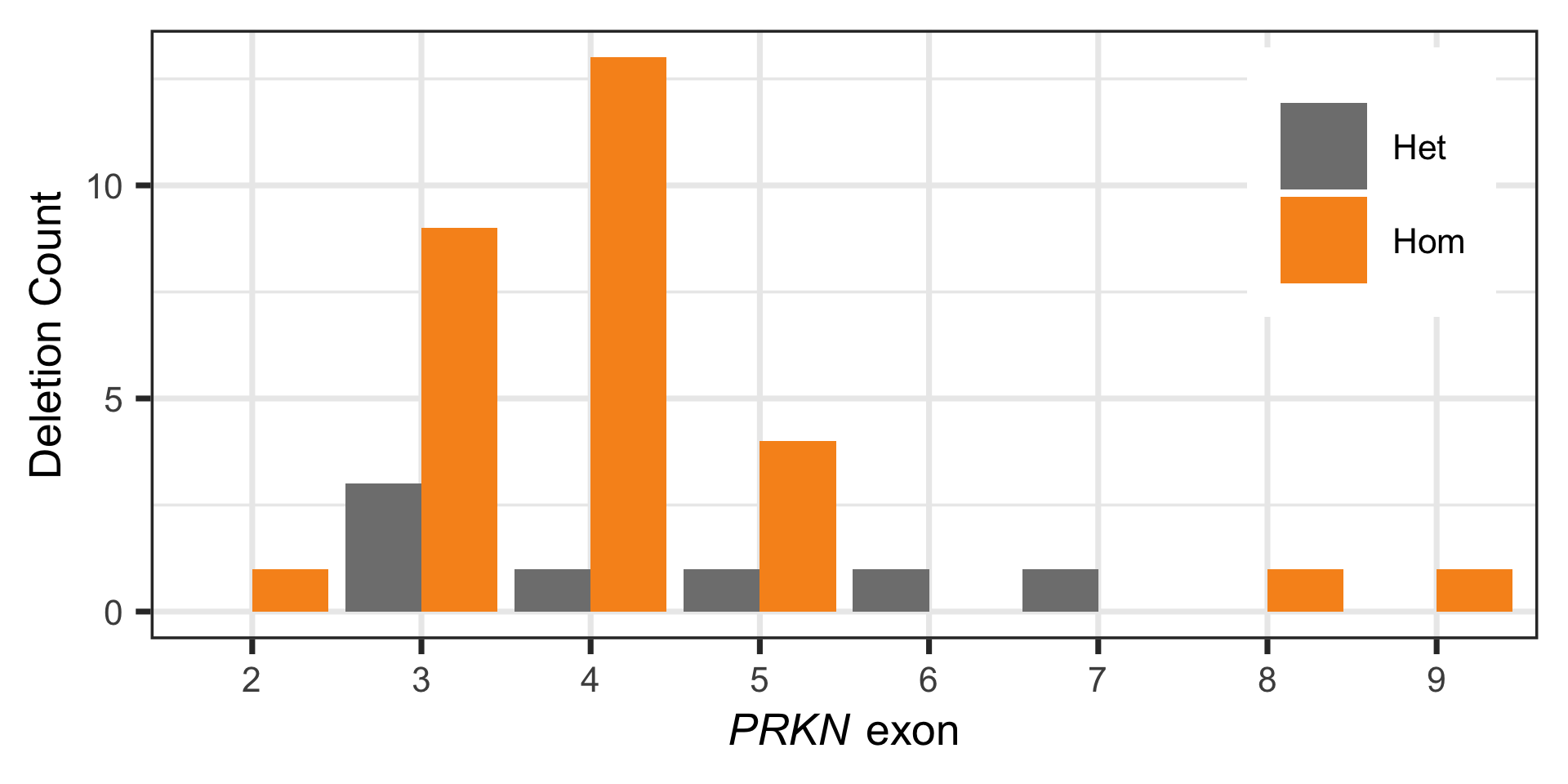

**Figure S10. A synonymous variant informed calibration procedure corrects platform differences between PD cases (WES data) and GAsPh2 controls (WGS data) for gene burden studies.** A) WES gene burden testing in synonymous variants only reveals inflation, indicating platform differences manifesting as case-control differences. B) Removing variants with differential coverage in cases and controls improves, but does not fully correct, inflation. C) Additional filtering of variants by dataset-specific variant quality thresholds (GATK parameter QualByDepth; QD) fully corrects inflation. D) Differential coverage and variant quality in case and controls for *DVL1*, whose gene burden p-values change significantly upon adjustment by coverage and variant-quality. See eMethods for description of λ_revised_ parameter and overall procedure.

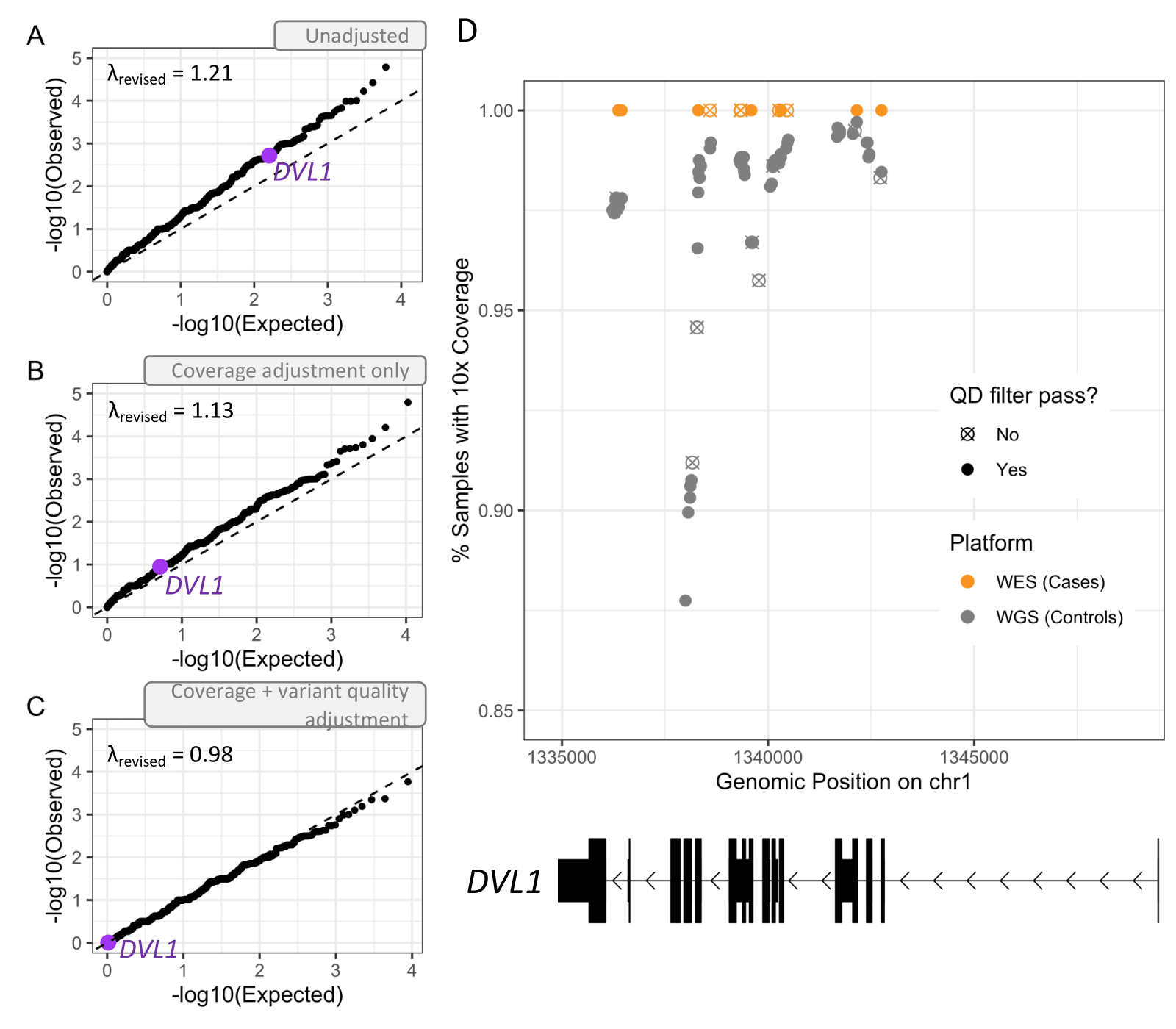

**Figure S11. Power curves for gene burden studies.** Estimates computed for 633 PD cases and 1,363 controls. Alpha set for 20,000 genes.

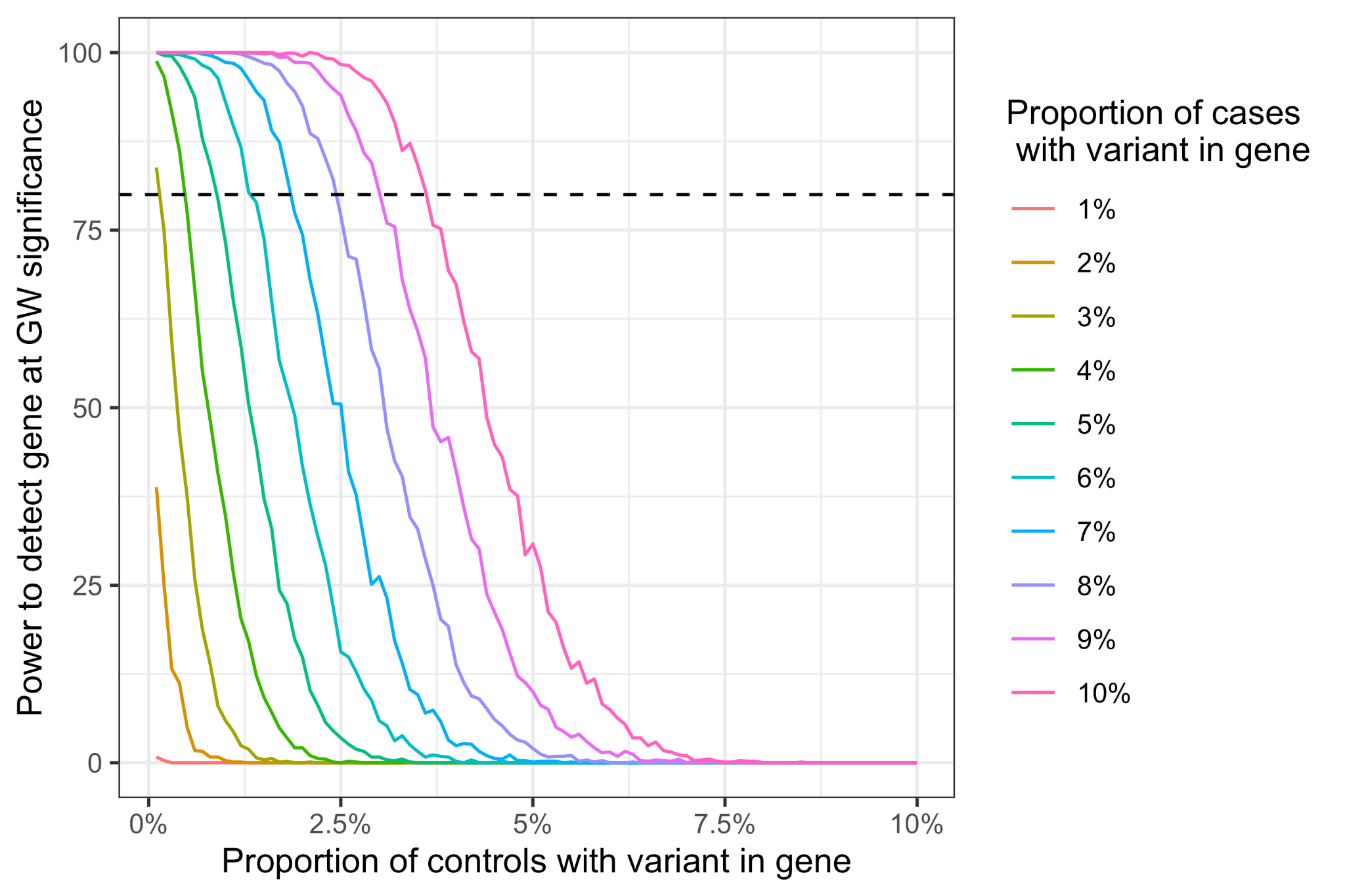

**Figure S12. BSN gene expression and variants driving differential gene burden result.** A) Tissue-level gene expression levels of BSN, derived from GTEx v8. B) List of *BSN* variants included under the qualifying variant criterion LoF + PolyPhen2 “probably damaging” or “possibly damaging” (p = 3.56E-4 in gene burden studies), along with their counts in cases and controls. Variants arranged by genomic/protein position.

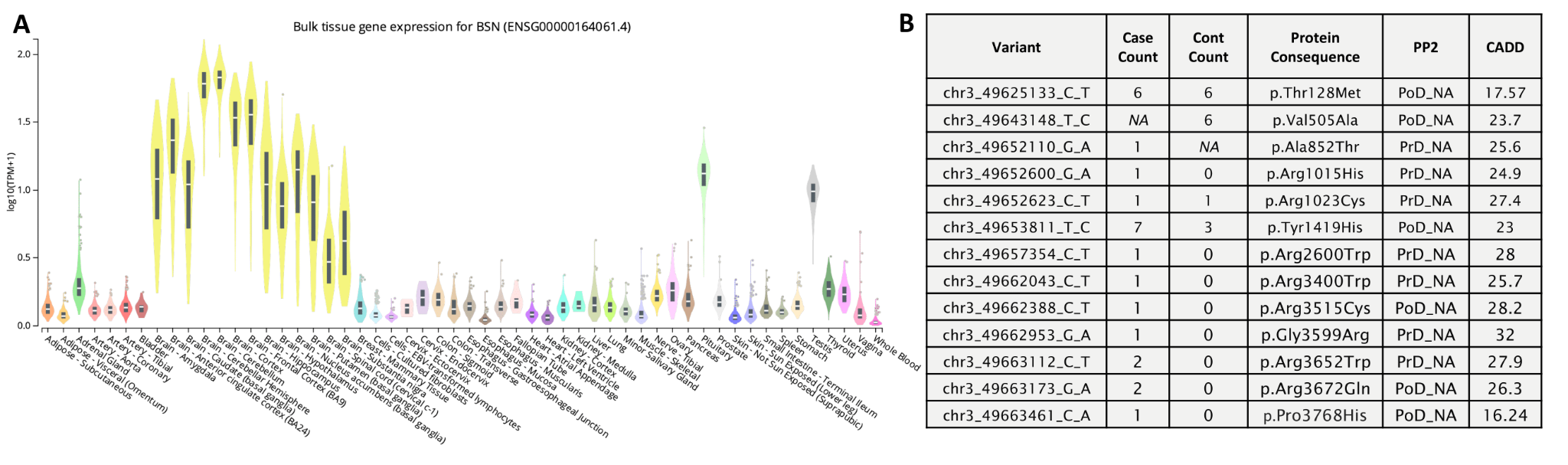
