## Supplementary Methods for "The genetic drivers of juvenile, young, and early-onset Parkinson’s Disease in India"

### **Supplemental Methods**

#### **Parkinson’s Disease cohort description and matched control selection**

Genetic data from a total of 674 Parkinson’s disease (PD) subjects were analyzed in this study. The subjects were recruited through a network of 10 specialty movement disorder centers and/or neurology clinics located across India. The study was approved by the Institutional Ethics Boards of each center, and written informed consent was obtained from each subject, as previously described^1^.

Across the 674 PD subjects, 21.1% (N = 142) were defined as familial PD (defined as ≥ 2 affected family members) and the remaining 78.9% (N = 532) were defined as sporadic PD, 69% (N = 466) were male, and the mean age of onset (AoO; available for N = 651 PD cases) was 39.3 years.

#### **Microarray genotyping & analysis of common variants**

##### **Genotype array data generation, processing, quality control, and imputation**

A total of 659 PD samples were profiled with the South Asian Research Genotyping Array for Medicine (SARGAM), a newly introduced genotyping array specifically designed for use in South Asian populations^2^. The raw data files (.CEL files) were analyzed with Axiom's APT command line best practices workflow, utilizing a DishQC cut-off of 0.82 and QC-call rate of 0.97. The QC-call rates are based on ~20,000 genomic markers, which are used to evaluate the genotyping quality of the sample. The plate-wise qc-call rate of the PASS sample average was above 98.5%. Further, joint genotyping was done on all samples available at the time of data analysis (N = 691; including additional samples not a part of this study) and yielded calls for 606,033 recommended markers out of 640,088 markers. Per the best practices workflow, recommended markers are defined as those classified as MonoHighResolution, PolyHighResolution, or NoMinorHom. Excluded markers are defined as those classified as CallRateBelowThreshold, OffTargetVariant, OtherMA, or Other.

After removing 7,572 markers that were mismeasured (see below), 598,461 markers across 691 samples were available from the SARGAM PD case data. Imputation, via Beagle 5.0^3^, was then performed using 6,461 samples from the GenomeAsia, Phase 2 (GAsPh2) reference panel, an extension of the previous GenomeAsia 100K project^4^. While the broader GenomeAsia effort sought to catalog Asian genomes as a whole, GAsPh2 was focused on developing a reference population for South Asian genomes specifically^2^. Imputation was performed separately for a merged PD case-control dataset (for use in PD diagnosis GWAS) and for a case-only dataset (for use in the age of onset GWAS). In the former instance, 516,388 autosomal markers were common between the SARGAM PD case and WGS GAsPh2 controls (see below).

Because the SARGAM array is a novel platform, we sought to investigate the frequency of genotyping errors of this new technology. We leveraged previously generated WGS data from 75 of the PD samples in our cohort (previously described^5^) as a gold standard against which the SARGAM variant calls (in these same samples) could be compared. First, we quantified between-platform allele frequency differences at all variants captured in both the SARGAM and WGS platforms and defined mismeasured variants as those with allele frequency differences > 1%. A total of 7,275 variants met this definition (Supplementary Fig. 3A).

Next, because the GAsPh2 control data were obtained via WGS, we hypothesized that variant calls mismeasured in PD samples from the SARGAM platform would tend to show significant p-values in a technology-agnostic case-control-based association study. To confirm this, we performed a GWAS of PD diagnosis (see additional details below) and regressed GWAS p-value against allele frequency difference. As expected, we observed that variants with lower p-values in the GWAS tended to have higher between-platform allele frequency differences (Supplementary Fig. 3B).

Because error from the mismeasured variants could be propagated to additional variants that were imputed based on those mismeasured variants, we removed the mismeasured variants from the dataset and repeated the imputation. We performed GWAS again in this new dataset, and regressed the new association p-values onto the between-platform allele frequency difference as measured in the *original* dataset. No association between GWAS p-value and (previous) between-platform allele frequency difference was observed (Supplementary Fig. 3C), confirming that we had successfully eliminated erroneous variant calls, and were no longer observing spurious associations .

Finally, we noticed many variants achieving genome-wide significance (*P* < 5E-8) in the PD diagnosis GWAS results only slightly exceeded a typically used threshold (Beagle DR2 = 0.8) to define high quality imputed variants (Supplementary Fig. 3D). Therefore, we elected to impose a more stringent imputation quality threshold (Beagle DR2 = 0.9), which resulted in a well-behaved QQ plot (Supplementary Fig. 3E).

##### **Ancestry-matched control selection**

Ancestry-matched controls were selected from the GAsPh2 population. GAsPh2 WGS data on 6,461 samples were restricted to SARGAM genomic positions captured on the SARGAM array and merged with SARGAM PD case data. Samples with a SNP call rate > 99.9% were retained, and variants were retained if they were autosomal, had a call rate > 95%, and had minor allele frequency (MAF) > 1%. The data were then LD pruned in PLINK 1.9^6^ with a window size of 50 Kb and an r^2^ value of 0.2, resulting in 276,081 markers in the merged dataset. Ancestry matching sample selection was done using UMAP with the following parameters: PCs = 15, Nearest neighbors = 15, Minimum distance = 0.5. With this approach, 1,376 control samples were selected. Finally, one member of 26 sample pairs demonstrating relatedness (via a PLINK 1.9^6^ “pi hat” metric > 0.2) was removed, leaving a final control population of 1,363 samples. Of these samples, the majority were male (N = 860; 63.1%) and the mean age was 54.5 years (interquartile range: 47 - 65 years).

As previously described^2^, the available GAsPh2 WGS data were obtained following GATK best practice QC recommendations, along with other additional stringent filters such as a variant quality score recalibration threshold of 99.9%. As a result, uniform marker coverage in the control population was not achieved, including a region containing the well-known PD gene *GBA*. This result is likely due to the established difficulty of sequencing this region, owing to the presence of a pseudogene (*GBAP1*) with a high degree of homology with *GBA*^7,8^. Therefore, case-control comparisons involving *GBA* variants were not possible in this study.

##### **Common variant (MAF > 5%) association studies and follow-up analyses**

To identify variants associated with PD diagnosis, a merged, imputed dataset consisting of SARGAM data from PD samples (N = 515, after restricting to those with sporadic PD and removing one sample from related pairs as described for controls) and WGS data from ancestry-matched controls derived from GAsPh2 (N = 1,363) was analyzed in PLINK 1.9^6^. The association testing model adjusted for sex, age, and the first 10 principal components to account for population stratification. After restricting to common variants (study MAF > 5%), 4,499,703 variants were included in the analysis.

We defined ranges for all loci reaching suggestive levels of significance (p < 1E-5) using the clumping command in PLINK 1.9^6^. For each of these loci, we tested for colocalization with signal from the most recent PD GWAS in European samples^9^ with the coloc R package^10^. For the identified signal in the *SNCA* region, we sought to nominate a causal gene for the signal using data that enabled variant-to-gene mapping and was relevant to the brain. We downloaded proximity ligation-assisted chromatin immunoprecipitation sequencing (PLAC-seq) data from sorted brain cell types^11^ and identified all regions across microglia, neurons, and oligodendrocytes that overlapped with any of the lead SNPs (p < 1E-5) to connect them to a gene promoter. Finally, we also compared effect sizes and p-values between our South Asian cohort and the same PD GWAS in Europeans for all variants included in both studies (n = 4,116,155 variants).

We also performed a case-only analysis to determine variants associated with AoO, restricting to sporadic samples with AoO < 50 in order to study age of onset of J/E/YOPD specifically, and removing one outlying sample with AoO = 3 (N = 484), again using PLINK v1.9. The association testing model adjusted for sex and the first 5 principal components. After restricting to common variants (study MAF > 5%), 4,246,454 variants were included in the analysis.

#### **Whole exome sequencing and analysis of rare variants**

##### **Whole exome sequencing data generation, processing and quality control**

To perform whole exome sequencing (WES) of PD samples, DNA was extracted from de-identified and consented patient blood samples (N = 576). Study subjects donated EDTA anticoagulated venous blood samples, and genomic DNA was isolated from whole blood by proteinase K digestion followed by phenol-chloroform extraction and precipitation in ethanol. Spectrophotometry was used to assess the quality and quantity of material. DNA concentrations were measured fluorometrically.

100ng of purified genomic DNA was subjected to mechanical fragmentation (Covaris) to obtain an average size of 200bp of DNA fragments. The fragmented DNA of each sample was subject to end repair, adenylation, adaptor ligation, and amplification to obtain whole genome libraries using the Kapa HTP library preparation kit (KAPA Biosystems, US). These libraries were hybridized with biotin-labeled whole exome capture probes present in the SureSelect Clinical Research Exome V2 (CREV2) whole exome panel (Agilent). The libraries were sequenced to a mean coverage > 80–100× with 150 nt paired-end reads on the Illumina sequencing platform (Hiseq2500 and HiSeqX, Illumina Inc).

Following quality check and adapter trimming using fastq-mcf (version 1.04.676), the sequencing reads were aligned to the human reference genome version GRCh38. The aligned reads were sorted, duplicate reads were removed, and the variants were called using the best practices pipeline of the Sentieon software (v201808.07). WGS from 92 PD samples (previously described^5^) were merged with WES dataset at the level of gVCF files to generate a jointVCF file, restricting to the CREV2 exome panel bed region. Variant-level QC was performed such as cluster size (window parameter =10), low quality (QUAL < 30.0), low overall depth (DP < 10) and low alt allele depth (AD < 7). In the sample-level filter, missing data rate was set at a threshold of 5%, although no samples were removed by this criteria. The median values for total aligned reads = 99.99%, reads above phred-scaled quality score of 30 (Q30) = 90.87%, and the duplicate reads = 8.03% (Supplementary Fig. 6) . For the WES data, the median panel on-target coverage was found to be 69.65%. The final WES dataset (defined henceforth as the merged WES and WGS PD case data) consisted of 718,181 variants and 668 samples.

##### **Diagnostic pipeline**

PD case WES data (N = 668) were used to evaluate the presence of pathogenic, likely pathogenic, or variants of uncertain significance (VUS) in known PD genes, as in our previous study^5^. Several variant annotation steps were undertaken to enable this analysis. First, gene annotation of WES variants was performed using the MedGenome in-house VariMAT tool, which uses the Variant Effect Predictor (VeP)^12^ program against the Ensembl release 91 human gene models^13^.

Next, the allele frequencies of detected variants in disease agnostic databases (gnomAD v3.0^14^, the Exome Aggregation Consortium (ExAC)^15^, 1000 Genomes^16^, and an internal MedGenome reference database of South Asian ancestry samples) was assessed. WES data variants were additionally annotated via *in silico* prediction tools, specifically Combined Annotation Depletion-Dependent (CADD) score^17^, Polymorphism Phenotyping v2 (PolyPhen2 or PP2)^18^, Sorting Tolerant from Intolerant (SIFT)^19^, MutationTaster2^20^, and LRT^21^, as well as disease databases, specifically the Online Mendelian Inheritance in Man (OMIM) database^22^, ClinVar^23^ and the Human Gene Mutation database (HGMD)^24^.

PD samples were ascertained for the presence of variants in a list of 64 PD genes, comprising three tiers of genes with differing evidence of a causal connection to PD. The first tier contained 13 very well characterized genes in PD. These genes have been reported in multiple independent families in literature, and were termed “primary” PD genes (see list below). The second tier contained 20 genes reported to be associated with PD but with “limited evidence”, i.e. reported in 1-2 families and/or not be replicated in independent datasets. The third tier contained 30 genes associated with “atypical Parkinson's or Parkinson's plus syndromes” , describing patients who present with Parkinson-like features.

- Tier 1 (“primary” PD genes) : DJ1, DNAJC6, FBXO7, PINK1, PLA2G6, PRKN, SYNJ1, VPS13C, ATP13A2, SNCA, CHCHD2, LRRK2, VPS35, GBA
- Tier 2 (limited Literature evidence for clinical diagnostics): ADORA1, PODXL, CSMD1, DNAJC13, TMEM230, LRP10, NR4A2, PANK2, PARL, PLXNA4 , RIC3, TNK2 , TNR, SNCAIP, CSF1R, EIF4G1, UCHL1, GIGYF2, HTRA2, NUS1
- Tier 3 (atypical Parkinson’s/Parkinson’s disease plus syndromes): ATP6AP2, C19orf12, GCH1, PRKRA, RAB39B, SLC30A10, SLC41A1, SLC6A3, SPR, TAF1, TRPM7, VAC14, ATP1A3, DCTN1, FTL, GRN, PNKD, PDYN, PRRT2, SGCE, SLC2A1, TOR1A, WDR45, PRKAR1B, PTRHD1, SPG11, ATXN2, TBP, ATXN8OS, MAPT

Samples were ascertained for variation within these genes, including single nucleotide variants (SNVs) and small indels, and copy number variants (CNVs) detected using the ExomeDepth (v1.1.10) method^25^. Common variants, defined as MAF > 5% in any population (annotated as described above) were excluded from the analysis. These PD gene variants were then assigned to classifications of pathogenic, likely pathogenic, or VUS taking into account variant zygosity and known mode of inheritance for each gene, based on American College of Medical Genetics (ACMG) criteria (Supplementary Fig.3).

Finally, samples were classified into several categories according to presence of these variants (groups of “Pathogenic”, “Likely pathogenic”, or “VUS”), or a “None” group consisting of samples lacking any rare variants having pathogenic, likely pathogenic, or VUS classification across all genes in the PD gene list. If a sample harbored multiple variants, they were assigned a mutation status according to the annotation of the most deleterious variant. For example, a sample with 1 pathogenic variant and 2 VUS variants was assigned to the “Pathogenic” group.

#### **GBA p.Ser164Arg Functional Characterization**

##### **Generation of p.Ser164Arg Knock-in (KI) cell line**

Homozygous knock-in of the GBA1-p.S164R variant was performed by Thermo Fisher Scientific (Carlsbad, CA) using CRISPR/Cas9 using Thermo Fisher’s optimized protocol. Briefly, cells were transfected with ribonucleoprotein (RNP) complexes of TrueCutTM Cas9 V2 + gRNA sequences, along with ss-Oligo donor repair templates containing the desired p.S164R edit. Two sequential rounds of transfection were utilized to increase editing efficiency for this cell line. Clonal populations of transfected cells were isolated via limiting dilution, and the resulting cells were screened for the GBA1-p.S164R knock-in edit via Sanger sequencing to identify clones with homozygous p.S164R SNP conversion. gRNA and ss-Oligo donor sequences used are provided in the table below.

| **Component** | **Sequence (5’ to 3’)** |
| --- | --- |
| gRNA | GAGAAGTCACAGCTGGCCAT |
| ss-Oligo donor (anti-sense strand) | GOFACTGGAAATCATCAGGGGTGTCTGCATAGGTGTAGGTGCGGATGGAGAAGTCACA  CCTGGCCATGGGTACCCGGATGATGTTATATCCGATTCCZFC |
| ss-Oligo donor (sense strand) | AZFTCTTGATCATCCTTTTCTGTAGGAATCGGATATAACATCATCCGGGTACCCATGGCC  AGGTGTGACTTCTCCATCCGCACCTACACCTATGCAGFOA |

##### **Cell lysis, immunoblotting, quantification of GCase protein levels**

Cells were lysed in RIPA buffer (Teknova, R3792) supplemented with an additional 1.8% SDS, cOmplete Protease Inhibitor Cocktail (Roche, #04693159001), PhosSTOP Phosphatase Inhibitor (Roche, #04906837001), and Benzonase nuclease (Sigma-Aldrich, E1014). Lysates were prepared for immunoblotting with NuPAGE LDS Sample Buffer (Thermo Fisher, NP0007) and NuPAGE Sample Reducing Agent (Thermo Fisher, NP0004), followed by incubation at 95 °C for 10 min to denature samples. Lysates were loaded onto 4-12% Bis-Tris NuPAGE gels (Thermo Fisher, WG1402) and fully resolved in 1x MOPS running buffer (Thermo Fisher, NP0001) before being transferred to a nitrocellulose membrane using the iBlot 3 Western Blot Transfer System (Thermo Fisher). Membranes were blocked with Intercept (TBS) Blocking Buffer (LI-COR, 927-60001) for 1 hour at room temperature, probed with rabbit anti-GBA (abcam, ab125065; 1/1000 dilution) and mouse anti-β-actin (Sigma-Aldrich, A2228; 1/2000 dilution) (diluted in Intercept Blocking Buffer + 0.2% Tween-20) overnight at 4ºC. Membranes were rinsed with 1x TBS-T, followed by incubation with IRDye® 800CW Goat Anti-Rabbit IgG (LI-COR, 926-32211) and IRDye® 680RD Goat anti-Mouse IgG (LI-COR, 926-68070), both diluted 1/10000, at room temperature for 1 hour. Membranes were rinsed 6 times, incubating 5 min at room temperature in every other wash, with TBS-T before imaging using the Odyssey CLx Infrared Imaging System (LI-COR). Band intensities were quantified using ImageStudioLite software (LI-COR). Band intensities were normalized to the median value within a replicate, and then GCase band intensity was normalized to the corresponding β-actin band intensity. The resulting ratio was then normalized to the ratio measured for the WT sample.

##### **Flow cytometric analysis of GCase activity**

A549 *GBA1* wild type, clonal *GBA1* knockout, and three clonal *GBA1* knockin S164R variant cell lines were grown in DMEM containing 4.5 g/L glucose and L-glutamine with 10% FBS at 37 °C and 5% CO_2_. Cells were seeded in 6-well plates (~100K cells per well) and grown overnight. The next day, cells were treated with 5 µM of the fluorescence quenched substrate LysoFQ-GBA^26^ at 37 °C for 3 hours. Cells were washed in PBS, lifted from plates with 0.05% trypsin-EDTA, and transferred to FACS tubes. Cells were then washed 2X in PBS and resuspended in PBS containing 3% FBS and 1% BSA. Cellular LysoFQ-GBA fluorescence was measured using the FITC channel on a BD FACSCanto II instrument equipped with a 488 nm laser and 530/30 filter set. At least 20,000 events were recorded per sample. Three biological replicates were performed, and median fluorescence intensity for each sample was calculated using FlowJo 10.9.0 software. For each sample within a biological replicate, the median FITC-A fluorescence intensity was normalized to the median of that biological replicate, followed by normalization to the *GBA1* wild type average. The scatter plot was generated in Prism version 10.0.2.

#### **Polygenic risk score analyses**

To derive a PD polygenic risk score (PRS), we used weights from 1,805 markers selected to define a PRS in a previous GWAS study of PD in Europeans^9^. Of these 1,805 markers, 1,354 markers were available in the imputed SARGAM case-control dataset and used to generate the PRS. We implemented an approach to estimate an ancestry-normalized PRS, allowing for the odds ratios to be implemented as weights in a different ancestry from which they were derived^27^.

A merged dataset of PD cases with available SARGAM data (who also had available diagnostic pipeline output; N = 624) and WGS controls (N = 1,363) was combined with an additional set of South Asian ancestry individuals (an internal MedGenome South Asian dataset), and a “raw” PRS was constructed with the European PD GWAS weights and allele count information at the corresponding variants. These PRS values were then regressed onto the first 5 principle components; the extracted residuals then comprised the ancestry-normalized PRS.

To test the association of the ancestry-normalized PRS with disease, decile cutoffs were defined only in the control population, and then all samples (cases and controls) were binned according to these deciles. Odds ratios for disease risk were calculated in each decile via a logistic regression model adjusted for age and sex, with the 5th and 6th decile groups together serving as a comparison group.

Finally, linear regression was used to compare the ancestry-normalized PRS values across controls and several diagnostic pipeline-defined groups for cases (“Pathogenic”, “Likely Pathogenic”, “VUS”, or “None”). Specifically, each PD case group was compared to controls, and then a within-case analysis was performed with the “None” group serving as a comparison group to the other 3 PD case classifications. Across all 7 performed tests, models included adjustments for age and sex.

#### **Gene-based burden testing of rare (MAF < 1%) variants**

We performed a gene burden analysis of rare variants, incorporating WES data from PD cases (N = 633, derived from N = 668 with WES data after removing one sample from related pairs as described above, N = 34, and PCA-based outliers, N = 1) and WGS data from controls (N = 1,363). Because case and control data had been ascertained using different technologies (WES and WGS, respectively) and had been preprocessed independently, we calibrated quality filters using synonymous variants, following an approach developed by Guo et al. for the analysis of a rare Mendelian disorder with public control data (specifically, gnomAD)^28^.

Guo et al. noted that synonymous variants are ideal for calibrating analysis settings (i.e. defining variant filters) across data sources because they are not expected to be associated with disease, particularly when aggregating them at the gene level. Therefore, any detectable association signal can be directly ascribed to technical artifacts. Variant filters include metrics of coverage and variant quality; as these metrics can be a function of allele frequency we only conducted calibration testing on rare synonymous variants (defined as MAF < 1% across any ancestral population as annotated by the VariMAT tool) since we sought to eventually test rare variants only. Finally, to account for potential differences in coverage and variant quality based on variant type, we restricted calibration and subsequent analyses to SNVs.

As a first step of implementing the calibration procedure, we conducted gene burden testing of rare, synonymous variants in the absence of variant filtering. Association statistics were calculated via a Fisher’s exact test derived from a 2x2 table comparing the number of samples with and without a qualifying variant in cases and controls. A dominant model was used such that any sample that had at least one qualifying variant in the gene was designated as a carrier. To quantify inflation in test statistics we calculated the lambda statistic defined by Guo et al. (“λ_Δ95_”). This metric, defined herein as λ_revised_, accounts for the large number of genes with p = 1, which itself results from small sample size and thus many genes with no qualifying variants. All subsequent gene burden testing, for both additional calibration testing and for the variants of interest, used these same approaches for burden testing and lambda estimation. We confirmed that gene burden testing of rare (MAF < 1%), synonymous variants, unadjusted for variant coverage and quality differences between cases and controls, resulted in significant inflation (λ_revised_ = 1.21; Supplementary Fig. 10A).

To adjust for read depth coverage differences between WES PD case and WGS control data, we applied a modification of the “Binomial Method” described by Guo et al. and originally proposed by Raghavan et al^29^. For each coding base, we compared the proportion of individuals covered at 5x, 10x, 15x, and 20x in the WES and WGS data via a binomial test. Sites that were not significantly different in coverage (p > 0.001) were retained for gene burden analyses. The coverage threshold that resulted in the least degree of inflation (determined via λ_revised_) was selected; in our cohort this threshold was 10x (λ_revised_ = 1.13; Supplementary Fig. 10B).

A final, subsequent variant filter was based on variant quality. We extracted the GATK QualByDepth metric (QD) for all rare SNVs in the WES and WGS data separately, and further determined dataset-specific values between the 60th and 95th percentiles in increments of 5. We filtered variants at every pairwise combination of these thresholds (again in WES and WGS data separately), performed gene burden testing, and selected the combination of thresholds that resulted in the least degree of inflation (determined via λ_revised_). In our cohort, the optimal thresholds were the 65th percentile in cases and 95th percentile in controls as imposing these parameters resulted in a full correction of inflation (λ_revised_ = 0.98; Supplementary Fig. 10C). For example, a strong disease association signal was observed in *DVL1* in the unadjusted analysis but was completely absent upon implementing selected coverage and variant quality thresholds (Supplementary Fig. 10D).

After selecting the coverage and (case and control dataset-specific) variant quality filters that best controlled Type I error in gene burden testing of rare, synonymous variants, we applied these same filters to all rare (MAF < 1% across any ancestral population; annotated as described above) SNVs in the dataset.

Next we implemented several different definitions of qualifying variants (QVs) across a spectrum of stringency. This approach, adapted from a recent gene burden study of Alzheimer’s Disease^30^, is a flexible strategy allowing for the investigation of the QV definition that provides the maximum evidence for differential burden, and for that definition to be gene-dependent. QV criteria were were defined as follows (from most stringent to least stringent): LoF variants only, LoF + PolyPhen2 (PP2) “probably damaging” variants, LoF + PP2 “probably damaging” or “possibly damaging” variants, CADD score > 20, and finally all missense, inframe, frameshift, and nonsense variants (a category referred to as “Functional Rare”).

Gene burden testing was conducted via the same methodology as described for the calibration procedure separately for each of these QV definitions. Therefore genes could be tested multiple times, with a maximum of 5 tests per gene. Multiple testing correction was applied across all tests, after implementing a minimum carrier count of 5 samples across both cases and controls.

Finally, to account for the potential for additional covariates to drive observed disease association, we did not perform tests for which there was a significant relationship between carrier status and gender (Fisher’s exact test p-value < 0.05), age (Wilcoxon test p-value < 0.05), or membership in the WGS subcohort of the PD case data (binomial test p-value < 0.05), examined for each gene-QV criteria combination. Power calculations were performed using the power.fisher.test function in the R package statmod.
